# Controlling the pandemic during the SARS-CoV-2 vaccination rollout: a modeling study

**DOI:** 10.1101/2021.03.24.21254188

**Authors:** João Viana, Christiaan H. van Dorp, Ana Nunes, Manuel C. Gomes, Michiel van Boven, Mirjam E. Kretzschmar, Marc Veldhoen, Ganna Rozhnova

## Abstract

There is a consensus that mass vaccination against SARS-CoV-2 will ultimately end the COVID-19 pandemic. However, it is not clear when and which control measures can be relaxed during the rollout of vaccination programmes. We investigate relaxation scenarios using an age-structured transmission model that has been fitted to age-specific seroprevalence data, hospital admissions, and projected vaccination coverage for Portugal. Our analyses suggest that the pressing need to restart socioeconomic activities could lead to new pandemic waves, and that substantial control efforts prove necessary throughout 2021. Using knowledge on control measures introduced in 2020, we anticipate that relaxing measures completely or to the extent as in autumn 2020 could launch a wave starting in April 2021. Additional waves could be prevented altogether if measures are relaxed as in summer 2020 or in a step-wise manner throughout 2021. We discuss at which point control of COVID-19 would be achieved for each scenario.

## Introduction

Mass vaccination against SARS-CoV-2 that started in Europe in December 2020/January 2021 [1] brings hope that the COVID-19 pandemic will end in 2021. Although the progress towards this goal is on the right track [2], many governments in Europe continue limiting socioeconomic activities to control the pandemic. Despite elaborate national vaccination schedules, it remains unclear when and which control measures can be relaxed and at which point the control of the pandemic will be achieved as the vaccination is rolled out in 2021. The understanding of how relaxation policies might affect the transmission of SARS-CoV-2 is further hampered by the emergence of novel variants [3, 4] that have a selective advantage, such as increased transmissibility [5–7] or the ability to reduce rapid neutralisation by the host [8]. For example, the current restrictions in Europe [9] are in part caused by a more transmissible [5–7] and potentially more pathogenic [10, 11] B.1.1.7 variant that originated in the UK and is quickly gaining dominance in other countries including Portugal [12, 13].

The vaccines that have been approved in Europe [14] show consistently high efficacy against severe disease, hospitalization and death in trials [15–17] and equally high effectiveness in real-world settings [18–22]. Multiple studies are under way to establish infection-blocking properties of these vaccines. Preliminary analyses of the national vaccination programme in Israel indicate that the effectiveness of the Pfizer-BioNTech vaccine against asymptomatic SARS-CoV-2 infections could be as high as 94% [21], as announced recently by the Israel Ministry of Health, Pfizer Inc and BioNTech SE. The recent Danish cohort study on long-term care facility residents and healthcare workers suggests that the effectiveness of the Pfizer-BioNTech vaccine based on a positive PCR test for SARS-CoV-2 is 64% and 90% beyond seven days of second dose in the two groups, respectively [19]. Similar results were found in a study among healthcare workers in England where the effectiveness of the Pfizer-BioNTech vaccine against symptomatic and asymptomatic infection was 86% seven days after two doses [22]. Based on the data from Israel, the effectiveness of the same vaccine against infection with SARS-CoV-2 was shown to be 51% 13-24 days after one dose [20]. Finally, in an analytical study by Lipsitch et al [23], the lower bound for the efficacy against transmission for one dose of Moderna vaccine was estimated at 61% but it could possibly be considerably higher, especially after two doses.

The consequences of relaxing control measures such as e.g., physical distancing, school closure, mask-wearing, test-and-trace and isolation, will depend on several factors, including the properties of vaccines deployed in a given country, specific vaccination schedules and speeds of vaccine rollout, but also the past epidemiology of SARS-CoV-2 that defines a proportion of the population protected after natural SARS-CoV-2 infection [24, 25]. All these factors are naturally country-dependent and will play a major role in how the pandemic will develop under different relaxation scenarios [26–29] and how quickly the control of COVID-19 will be gained in specific countries throughout 2021 and possibly beyond. To make a few distinctive examples, we recall Israel which has the highest vaccination rate worldwide so that, on average, every person has received at least one vaccine dose by mid-March 2021 [1] and Manaus in Brazil, where the levels of protection by natural infection close to the theoretical herd immunity threshold were achieved prior to the start of mass vaccination [30].

An extensive body of literature addresses the challenges of modelling real-time fast-moving COVID-19 pandemic [31]. Mathematical transmission models robustly calibrated to available data are among the best tools available to provide input into the discussion on the response to the COVID-19 pandemic [32–43] and they will continue to play an important role in making decisions surrounding the relaxation of measures in 2021 [26–29]. Several modeling studies provided support for the development of COVID-19 vaccines and early planning of vaccination scenarios and rollouts [44–48], but these models typically assumed that a large proportion of the population is vaccinated instantaneously and/or did not focus on relaxation strategies. More recently, organized teams of modeling experts supporting decision-makers over health emergencies in China and the UK evaluated the roadmap scenarios for relaxation of control measures in these countries in light of ongoing mass vaccination [26–28].

The present study makes a contribution towards better understanding of when and which control measures can be relaxed as mass vaccination programmes progress in 2021. We take Portugal as a case study where good quality data for model parameterization are available but, apart from efforts of genomic surveillance [49], there are no dedicated COVID-19 modeling studies for informing policymaking in this country [50]. Using an age-structured transmission model that has been fitted in a Bayesian framework to the data from various sources (age-specific hospitalizations and seroprevalence, social contact and demographic data, national vaccination plan and vaccine rollout data etc.), we investigate future pandemic trajectories under several alternative relaxation scenarios throughout 2021. Among the explored relaxation strategies are lifting measures to the same extent as in summer 2020 and later on in autumn 2020, the complete lifting of measures and combinations of these. We evaluate the impact of each scenario on the epidemic dynamics as quantified by projected hospital admissions, the time-dependent effective reproduction number, population immunization level due to natural infection and vaccination, and timing of reaching control of COVID-19 in Portugal. Finally, we discuss the implications of our findings for the post-pandemic dynamics of SARS-CoV-2.

## Results

### Model calibration

The model was fitted to age-stratified COVID-19 hospitalization data in the period from 26 February 2020 till 15 January 2021 and cross-sectional age-stratified SARS-CoV-2 seroprevalence data assessed from 21 May 2020 till 8 July 2020. The model reproduces well the age-specific hospital admissions (Figure 1) featuring (i) the first pandemic wave (March-April 2020), (ii) relatively low epidemic activity (May-August 2020), (iii) the second pandemic wave (September-mid-December 2020), (iv) the third wave that started in mid-December 2020 and was still ongoing on 15 January 2021 [51]. The estimated hospitalization rates increase with age from 0.12 (95%CrI 0.07-0.23) per year for children under 5 years of age to 14.24 (95%CrI 9.91–21.23) per year for persons above 80 years (Figure S1). In agreement with other studies [52, 53], the estimated susceptibility to SARS-CoV-2 increases with age (Figure S2). The meaning of model parameters is given in Tables S2 and S4, and their estimates are shown in Figures S1 and S2.

**Figure 1.**
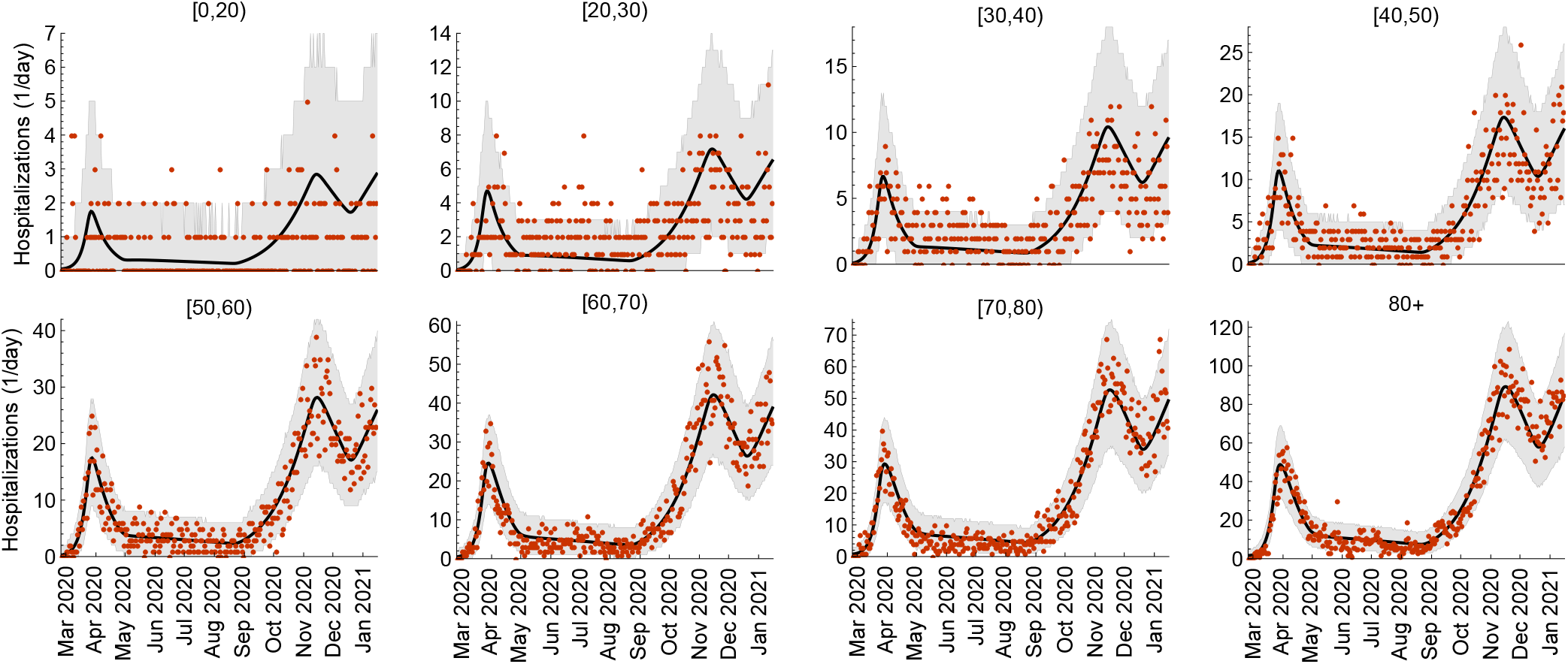
Model fit to COVID-19 hospitalizations. The age-stratified daily hospital admission data are shown as red dots. The median trajectories estimated from the model are shown as the black lines. The gray shaded regions correspond to 95% Bayesian prediction intervals based on 2,000 parameter samples from the posterior distribution. Hospital admissions were estimated for 10 age groups (see Methods). For presentation purposes, here we grouped hospitalizations for ages [0,5), [5,10), [10,20).

The model also reproduces well the age-specific and total seroprevalence in the population (Figure 2). The estimated age-specific seroprevalence ranged between 1.77% (95%CrI 0.98–2.91%) for 1 to 10 years old children to 4.61% (95%CrI 3.47–5.91%) for 20 to 40 years old adults (Figure 2 **a**). The total seroprevalence steadily increased with time reaching 19.37% (95%CrI 14.82–24.57%) on 15 January 2021 (Figure 2 **b**).

**Figure 2.**
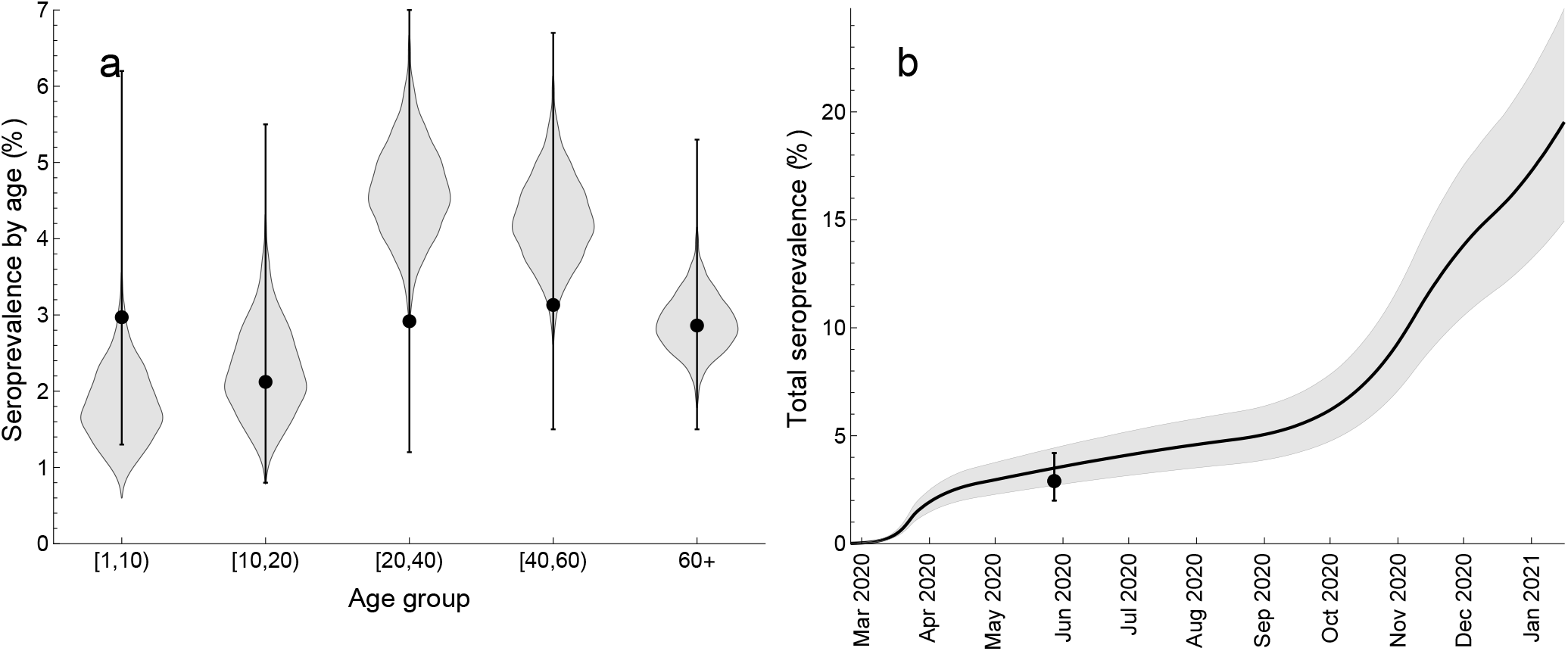
Model fit to SARS-CoV-2 seroprevalence. **a** Age-specific seroprevalence. **b** Total seroprevalence. The data (dots and error bars) are based on the cross-sectional seroepidemiological survey (First National Serological Survey) conducted after the first pandemic wave [54]. **a** The violin shapes represent the marginal posterior distribution of the age-specific seroprevalence in the model. **b** The black line and the gray shaded region show the median total seroprevalence and 95% credible intervals. The uncertainty in the model is based on 2,000 parameter samples from the posterior distribution. The total seroprevalence refers to population older than 1 year [54].

### Time-varying contact patterns and effective reproduction number

We estimated how age-specific contact rates in the population changed due to control measures as the pandemic developed. These contact rates denote the average number of transmission-relevant contacts per day a person in a given age category has with persons in other age categories. We further calculated the time-dependent effective reproduction number, *R*_*e*_(*t*), defined as the average number of secondary infections caused by one infectious individual in the population with age-specific contact patterns and age-specific seroprevalence at time *t. R*_*e*_(*t*) < 1 signifies the control of the pandemic with possibly some of control measures in place. The full control of COVID-19 is achieved when *R*_*e*_(*t*) < 1 and the contact rates in the population are restored to the pre-pandemic level.

Our findings are summarized in Figure 3, where we show the total daily hospitalizations (Figure 3 **a**), the average (over all ages) number of daily contacts in the population (Figure 3 **b**) and *R*_*e*_(*t*) (Figure 3 **c**) evaluated bi-weekly in the period from 26 February 2020 till 15 January 2021. The green vertical lines indicate the estimated mid-point transitions in the age-specific contact rates (see Methods). The pre-pandemic average number of daily contacts was 12.6. The estimated basic reproduction number (in the absence of control measures and with zero seroprevalence) was 2.20 (95%CrI 1.97–2.56). The control measures introduced during the first wave in spring 2020 reduced the number of contacts to 4.2 (95%CrI 3.3–5.0) and *R*_*e*_ to 0.69 (95%CrI 0.64—0.75). After some of these measures were lifted, the number of contacts increased to 5.9 (95%CrI 5.1–6.6) and *R*_*e*_ increased to almost 1 and stayed nearly constant throughout summer 2020. At the start of the second wave in autumn 2020 that followed the opening of schools and the associated changes in the contact patterns of the rest of the population, the average number of contacts further increased to about 7.6 (95%CrI 6.7–8.3) and *R*_*e*_ to 1.24 (95%CrI 1.21–1.28). The reinforcement of measures during the second wave could only reduce *R*_*e*_ to 0.89 (95%CrI 0.86–0.99) as compared to *R*_*e*_ of 0.69 after more severe measures introduced during the first wave. Finally, the increased activity of the population around Christmas and the New Year 2021 initiated the third wave in January 2021.

**Figure 3.**
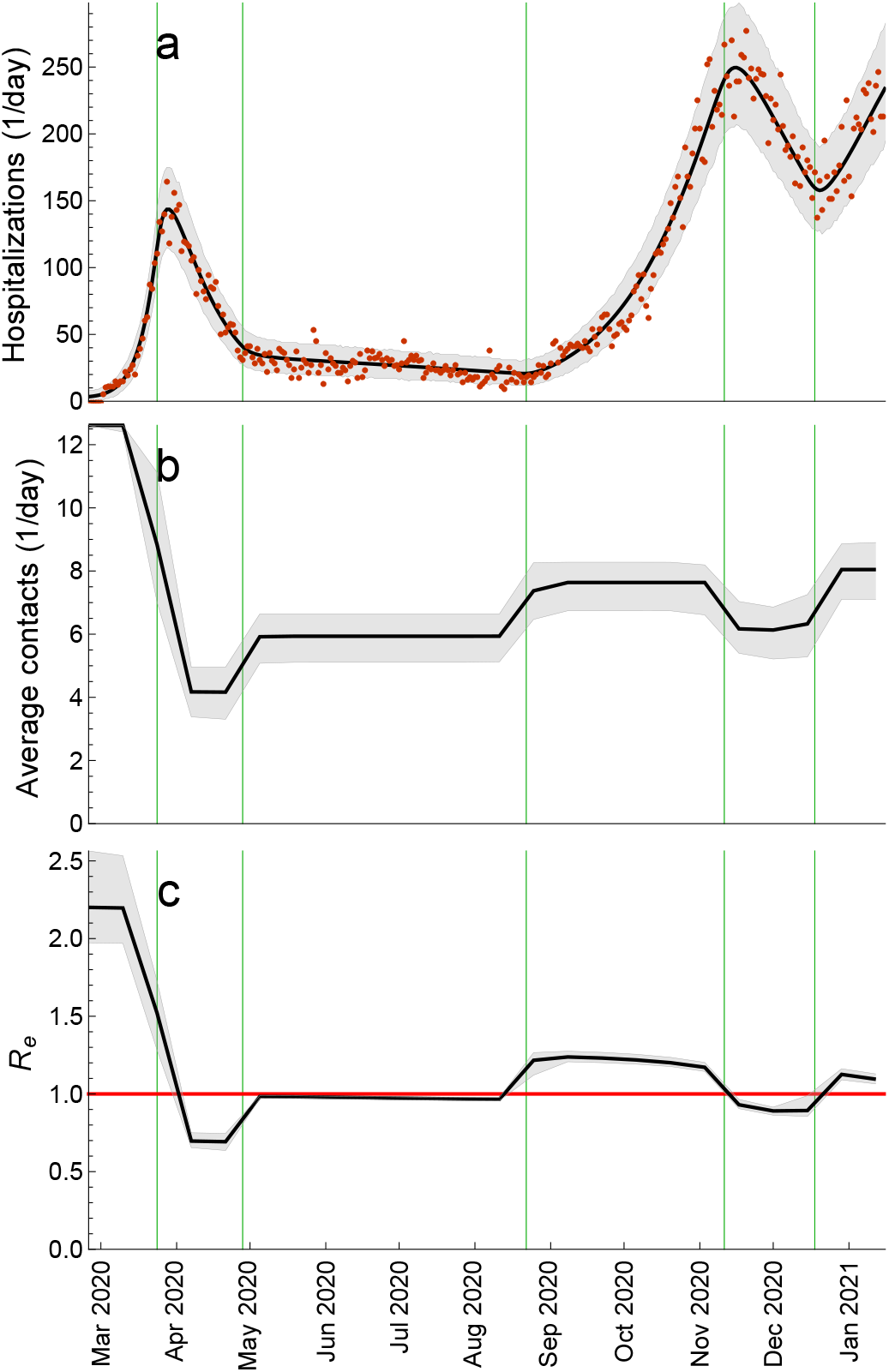
Estimated contact rate and effective reproduction number. **a** Total daily hospital admissions with COVID-19. **b** Average (over all ages) number of daily contacts in the population. **c** Effective reproduction number, *R*_*e*_(*t*). The average daily contacts and *R*_*e*_ were evaluated once every two weeks. The green vertical lines indicate the estimated mid-point transitions in the age-specific contact rates. The red horizontal line denotes *R*_*e*_ = 1. The hospitalization data are shown as red dots. The black solid lines are the median trajectories estimated from the model. The gray shaded regions correspond to 95% credible intervals.

### Vaccination rollout

We implemented the rollout of vaccination against SARS-CoV-2 as set out by the Directorate-General of Health — a division of Portuguese Ministry of Health concerned with public health (Table 1). The mass vaccination started on 27 December 2020, is planned to proceed in three phases that will cover the whole population of Portugal by 31 December 2021. In the model, we made several simplifying assumptions regarding vaccination, i.e. 1) at most 90% of each age group will be vaccinated (as supported by the survey conducted between 23 January and 5 February 2021 on the willingness to get vaccinated where the percentage of the Portuguese residents who want to get vaccinated exceeds 95% [55]) except for persons under 20 years of age (as supported by the current guidelines on the ineligibility for vaccination of persons under 18 years of age); 2) the distributed vaccine is by BioNTech/Pfizer brand (as supported by the recent ECDC vaccination data for Portugal where 96% of vaccine doses distributed up until February 21, 2021 are by BioNTech/Pfizer); 3) vaccination is modelled as a single event that immediately confers protection equivalent to two vaccine doses; 4) we considered an infection-blocking vaccine and formulated optimistic (main results) and pessimistic (sensitivity analyses) assumptions for vaccine efficacies in reducing infection, disease and severe disease; 5) there is no waning of protection against (re-)infection after natural infection and vaccination. More details of the vaccination model are given in Methods.

**Table 1.** The Portuguese vaccination plan.

| Category | Age (years) | Vaccination period | Persons |
| --- | --- | --- | --- |
| **Phase 1** |  |  | **937,361** |
| Healthcare workers (HCW) | 20 – 65 | 27 Dec 2020 – 28 Feb 2021 | 199,708 |
| Long-term care facilities (LTCF) |  | 01 Jan 2021 – 28 Feb 2021 | 148,119 |
| Residents | 65+ |  | 86,982 |
| Staff | 20 – 65 |  | 61,138 |
| Risk Group 1 | 50+ | 01 Feb 2021 – 30 Apr 2021 | 513,634 |
| Cardiac insufficiency |  |  | 207,571 |
| Coronary heart disease |  |  | 169,265 |
| Renal insufficiency |  |  | 8,201 |
| Chronic obstructive pulmonary disease (COPD) |  |  | 128,597 |
| First response professionals (FRP) (firemen, police, military etc.) | 20 – 65 | 01 Feb 2021 – 30 Apr 2021 | 75,900 |
| **Phase 2** |  |  | **3,333,191** |
| Persons with or without morbidities unvaccinated before* | 65+ | 01 May 2021 – 31 Jul 2021 | 1,873,349 |
| Risk Group 2 | 50 – 65 | 01 May 2021 – 31 Jul 2021 | 1,459,842 |
| Diabetes |  |  | 222,864 |
| Neoplasm |  |  | 114,246 |
| Hepatic insufficiency |  |  | 93,004 |
| Chronic kidney disease |  |  | 4,222 |
| Obesity |  |  | 392,959 |
| High blood pressure |  |  | 632,547 |
| **Phase 3** |  |  | **6,529,448** |
| Remaining persons (excluding children)** | 20 – 65 | 01 Aug 2021 – 31 Dec 2021 | 6,529,448 |
| **Total\*** |  |  | **10,800,000** |

\*The Portuguese vaccination plan assumes that all persons in the population will be vaccinated with a two-dose vaccine schedule. In the model, the maximum vaccination coverage in any age group is 90%. \*\*According to the current guidelines, persons under 18 years old are not eligible for vaccination. In the model, we assumed that the age group of 0 to 20 years old is not vaccinated.

We used the rollout schedule (Table 1) and data (Figure 4 **a**) on the age distribution of morbidities among the Portuguese residents and age distribution of prioritized vaccination categories (e.g., healthcare workers, long-termM care facilities staff and residents etc.) to calculate age-specific vaccination rates (number of persons in a given age group vaccinated per day) as the vaccination programme progresses (Figure 4 **b**; see Figure S3 for detailed information). The vaccination rate refers to vaccination with two vaccine doses. The maximum vaccination coverage of 90% is projected to be reached in the following order (Figure 5 **a**): 80+ (29 June 2021), [60,80) (20 July–23 July 2021), [50,60) (29 August 2021) and [20,50) (16 November 2021) (see Figure S4 for absolute numbers of vaccinated persons). The total coverage in the population will increase to 9%/38%/73% (maximum coverage) by 1 May/1 August/16 November 2021 (Figure 4 **b**). The ECDC vaccination rollout data for Portugal agree well with these projections.

**Figure 4.**
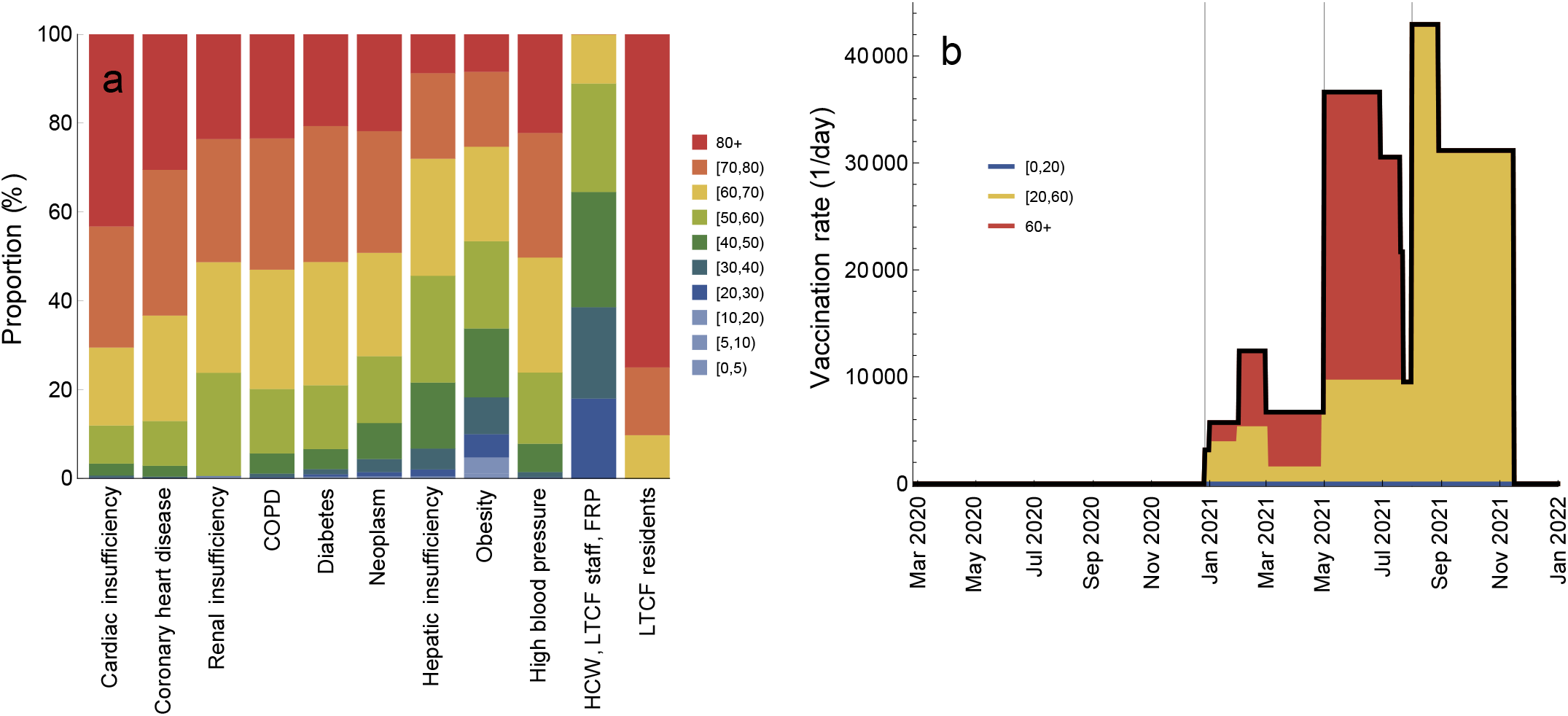
Vaccination rollout schedule. **a** Age distribution of vaccination categories. **b** Total vaccination rate (number of persons vaccinated per day, black line) and proportions of vaccination rate attributable to ages [0,20) (blue), [20,60) (yellow) and 60+ (red). The gray vertical lines in **b** indicate the starting dates for different vaccination phases (Table 1). The age-specific vaccination rates are given in Figure S3.

**Figure 5.**
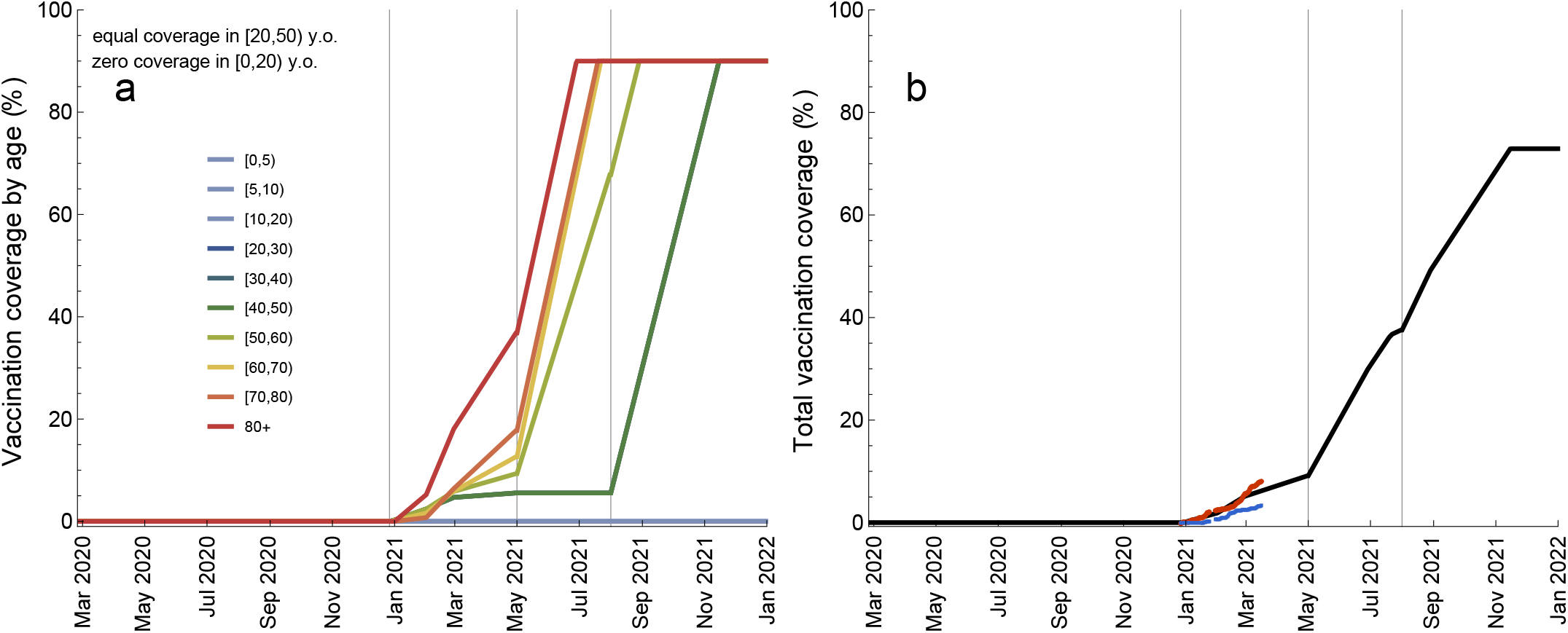
Vaccination coverage during the vaccination rollout. **a** Age-specific coverage (percentage of vaccinated persons per age group). **b** Total vaccination coverage (percentage of vaccinated persons in the population). The gray vertical lines indicate the starting dates for different vaccination phases (Table 1). The coverages for ages [20,30), [30,40), and [40,50) are equal (see Figure S4 for the absolute numbers of vaccinated persons). The coverage for ages [0,20) is zero. The ECDC vaccination rollout data in **b** are shown as red (1 dose) and blue (2 doses) dots.

### Scenarios for relaxation of control measures

To account for the epidemiological situation in Portugal between mid-January and mid-March 2021 [51], we modeled the third wave of hospitalizations that was curbed by the substantial reinforcement of measures similar to those implemented during the first wave in spring 2020. We also modelled an increase in the transmisibility of the virus due to the rapid spread of B.1.1.7. variant in Portugal. The situation in mid-march 2021 is then described by the average number of daily contacts of 4.2, *R*_*e*_ of 0.67 and the circulating variant that is 50% more transmissible [5–7] than the original variant that was dominant in Portugal until December 2020. Starting from this situation, we generated scenarios for relaxation of control measures as follows (Figure 6): Scenario 1) lifting all measures so that contact rates in the population return to the pre-pandemic level (average rate of 12.6 contacts/day); Scenario 2) partial lifting of measures that increases contact rates to the level of September-October 2020 (7.6 contacts/day); Scenario 3) partial lifting of measures that increases contact rates to the level of June-August 2020 (5.9 contacts/day). In accordance with the plan of the Portuguese government to alleviate some of the current measures in spring 2021 and to make the scenarios comparable, we used the same mid-point (1 April 2021) and the same speed of transition between the contact levels (10 days).

**Figure 6.**
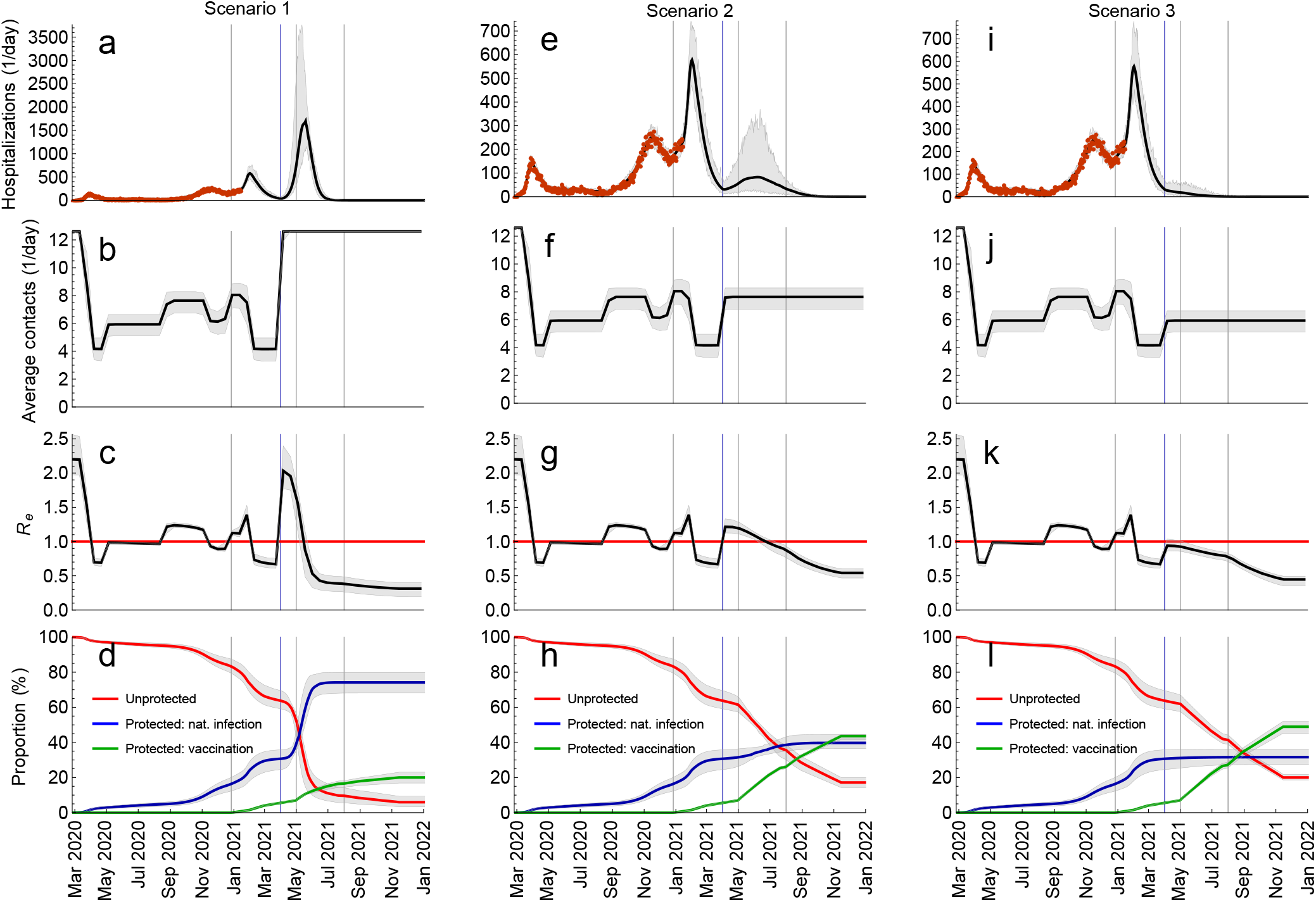
Scenarios for relaxation of control measures. **a**-**d** Lifting all measures so that contact rates in the population return to the pre-pandemic level. **e**-**h** Partial lifting of measures so that contact rates increase to the level of September-October 2020. **i**-**l** Partial lifting of measures so that contact rates increase to the level of June-August 2020. The blue vertical lines indicate the mid-point of the transition (1 April 2020). The gray vertical lines indicate the starting dates for different vaccination phases (Table 1). The red horizontal line denotes *R*_*e*_ = 1. The hospitalization data are shown as red dots. The thick solid lines are the median trajectories estimated from the model. The gray shaded regions correspond to 95% credible intervals.

The comparative analysis of Scenarios 1, 2, and 3 is shown in Figure 6. The model predicts that lifting all measures (Scenario 1; Figure 6 **a**-**d**) launches a fourth wave that is significantly larger than the previous waves, resulting in 58,226 cumulative hospitalizations between 1 April and 1 January 2022 (Figure 6 **a**). *R*_*e*_ increases sharply from 0.67 on 23 March 2021 to 2.03 two weeks later (Figure 6 **c**) which is very close to the basic reproduction number of 2.20 at the start of the pandemic. The full control over COVID-19 is reached on 18 May 2021 when *R*_*e*_ drops below 1 and the contact rates are the pre-pandemic level (Figure 6 **b**). At this threshold, 60% of the population acquired protection after natural infection and only 10% are protected after vaccination (Figure 6 **d**). Relaxing measures according to Scenario 2 (Figure 6 **e**-**h**) initiates a new pandemic wave too, albeit smaller in magnitude than Scenario 1 (8,975 hospitalizations between 1 April and 1 January 2022; Figure 6 **e**). In this case, *R*_*e*_ becomes smaller than 1 on 29 June 2021 (Figure 6 **g**) but the measures have to be kept in place (Figure 6 **f**) to control the spread. The increase of contact rates to the level of June-August 2020 (Scenario 3; Figure 6 **i**-**l**), however, does not lead to a rise in hospitalizations (Figure 6 **i**) because *R*_*e*_ stays below 1 (Figure 6 **k**) but, like in Scenario 2, the measures have to continue until sufficient number of people acquire protection by vaccination to relax them completely. In addition, we explored Scenario 4 (Figure 7) where measures are relaxed in a step-wise manner so that contact rates first rise to the level of June-August 2020 (Step 1, Scenario 3), then to the level of September-October 2020 (Step 2, Scenario 2) and, finally, to the pre-pandemic level (Step 3, Scenario 1) (Figure 7 **b**). The mid-points of transitions were 1 April, 1 June and 1 October 2021 (blue vertical lines in Figure 7) and the relaxation speed of 10 days was used for all transitions. In this scenario, additional waves can be prevented altogether and hospitalizations stay at the level comparable to that in summer 2020 when the epidemic activity was low (Figure 7 **a**). Interestingly, Step 2 (1 June) and Step 3 (1 October) increase *R*_*e*_ above 1 (Figure 7 **c**) leading to waves of infections (Figure S5) but a large increase in hospitalizations is not observed because a substantial proportion of the vulnerable population has been vaccinated (Figure 5). The full control of the pandemic (*R*_*e*_(*t*) < 1 and pre-pandemic contact rates) is reached on 8 February 2022 (Figure 7 **c**) when 36% of the population are protected after natural infection, 48% after vaccination, and 17% stay unprotected (Figure 7 **d**). This is drastically different from Scenario 1, where the control was reached mainly due to protection through natural infection (60%), and the minority was protected by vaccination (10%).

**Figure 7.**
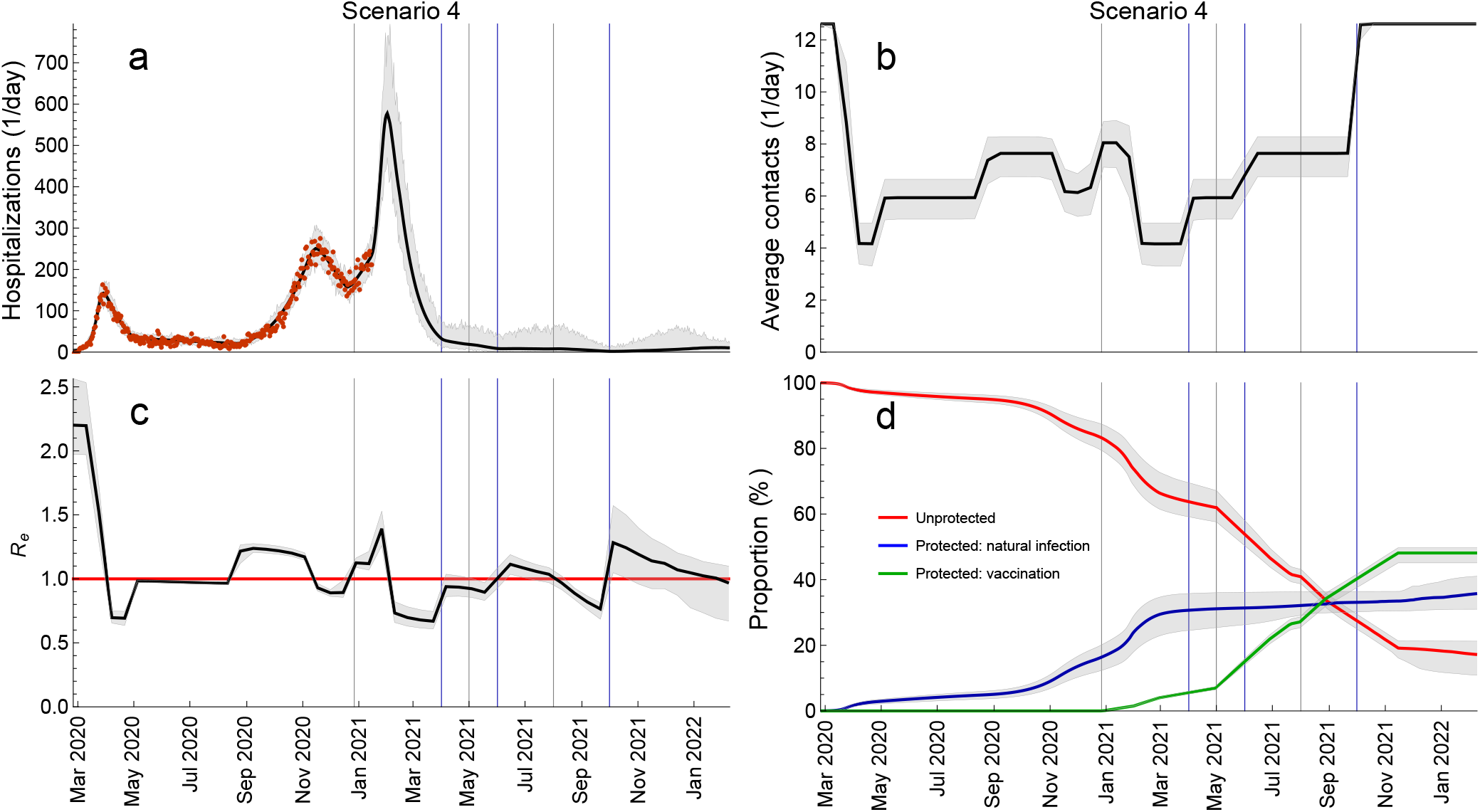
Sequential relaxation of control measures. This scenario consists of sequential relaxation of measures so that the contact rates increase, in sequence, to the level of June-August 2020, of September-October 2020 and the pre-pandemic level. The blue vertical lines indicate the mid-points of these transitions (1 April, 1 June, 1 October). The gray vertical lines indicate the starting dates for different vaccination phases (Table 1). The red horizontal line denotes *R*_*e*_ = 1. The hospitalization data are shown as red dots. The thick solid lines are the median trajectories estimated from the model. The gray shaded regions correspond to 95% credible intervals.

Finally, we would like to stress that for demonstration purposes the timings of Steps 2 and 3 in Scenario 4 have been intentionally chosen so that the epidemic activity (i.e., the number of hospital admissions) in 2021 is similar to that in summer 2020. The premature relaxation of measures can still lead to new waves of hospitalizations. We demonstrate this in Figure S6 where Step 3 occurs on 1 August instead of 1 October 2021. Similarly, the results presented for Scenario 4 (and other scenarios as well) are the most optimistic in terms of projected hospitalizations and get worse for a pessimistic set of vaccine efficacies or if individuals return to pre-pandemic contact rates immediately upon getting vaccinated (see Figure S7).

## Discussion

In this study, we used an age-structured model for SARS-CoV-2 transmission to generate several scenarios for relaxation of control measures during the ongoing vaccination rollout in Portugal. In agreement with the plans of the Portuguese government, the mid-point of easing of measures is April 2021. Our analyses demonstrate that vaccination alone, if rolled out according to the national vaccination schedule, is likely to be insufficient to control the Portuguese pandemic when control measures are significantly alleviated in April 2021. Returning to the prepandemic lifestyle already in spring 2021 is the worst-case scenario that would be detrimental for the healthcare system. Even for the most optimistic model assumptions, this scenario would result in a wave of hospitalizations several orders of magnitude larger than the three previous waves. Relaxing measures to the same extent as in autumn 2020 would lead to somewhat smaller wave (as compared to the worst-case scenario and even to the third wave that actually occurred) that would, nonetheless, present a significant burden for the national healthcare. These findings are qualitatively similar to those in modeling studies for China [28] and the UK [26, 27], but the quantitative comparison is not possible because of different settings and contexts in which those studies were conducted. Additional waves could be prevented altogether if measures in spring 2021 are relaxed to the same extent as in summer 2020 or in a step-wise manner throughout 2021.

The point at which the pandemic is brought under full control (*R*_*e*_(*t*) < 1 and pre-pandemic contact patterns) depends on the amount of protection in the population acquired through a combination of natural infection and vaccination. Gaining the control quickly (by mid-May 2021) occurs mainly through protection by natural infection (60% of the population) while the minority (10%) would be protected by vaccination. As mentioned above, this worst-case scenario is, obviously, undesirable and is not very much different from letting the pandemic develop without any control measures. In the gradual relaxation scenario, achieving control takes more than one year since the start of vaccination rollout, but almost 50% of the population are protected by vaccination and a smaller proportion (35%) have experienced SARS-CoV-2 by that point. Alternative to these scenarios would be accelerating the vaccination campaign so that vaccination coverage increases faster than initially projected and confirmed by the ECDC vaccination rollout data [2]. However, it is not clear whether this option is viable for Portugal given the current shortage for COVID-19 vaccines.

A strength of our analyses is that we calculate the effective reproduction number using the estimated current levels of age-specific seroprevalence and vaccination coverage in the population instead of reducing the value of *R*_*e*_ at the beginning of the pandemic homogeneously across age groups as it is done in e.g. the study for China [28]. Another strength is that, unlike this study [28] and the studies for the UK [26, 27], the parameters of our model are statistically evaluated to match the course of the Portuguese pandemic as reflected by age-specific hospital admissions and age-specific seroprevalence [54]. In addition, our fitting procedure allows for estimation of temporal changes in age-dependent contact patterns as a response to prior control measures during this pandemic. Therefore, instead of modeling specific relaxation policies, that are notoriously hard to implement in mechanistic transmission models, we model several scenarios using the estimated contact structure after relaxation of measures in summer and autumn 2020.

In light of these past measures, our findings are easy to interpret and contain an important message for local policymakers. School opening is thought to be the main driver of the changes observed in autumn 2020, although an increase in socializing indoors in general caused by weather alone must also have played a role. If the relaxation planned for April 2021 includes school reopening in full after Easter and resuming indoor service in restaurants and bars, then it is very likely that the average contact rate in the population will reach levels very similar to those in autumn 2020. As a consequence, this might lead to a new wave of hospitalizations as illustrated in Scenario 2. On the bright side, according to our analysis the goal of Scenario 3, in which major waves are avoided, seems well within reach, given the light control measures that were in place during summer 2020. Combining these with some additional limitations of indoor social activities and online classes for secondary school students could help to replicate the average contact rate of summer 2020, compensating for opening of elementary schools.

As any model, our model has limitations. An important one is that protection against (re-)infection after natural infection and vaccination is permanent over the time-scale of our analyses (almost two years). This frequently used assumption [26–28, 44, 47] leads to that in our model, theoretically, SARS-CoV-2 can be eliminated from the population. However, as we discussed recently [56] and as addressed in several conceptual modeling studies [57–59], accumulating evidence suggests that after the initial pandemic phase SARS-CoV-2 is likely to be transitioning to endemicity and continued circulation. Specifically, recent data from individual-level studies point to that detectable levels of antibodies to SARS-CoV-2 providing immunity against reinfection can wane on the time scale of a few months to few years following exposure, as shown by our group [60] and corroborated with findings of other studies [61–63]. However, the immunity to SARS-CoV-2 depending on a combination of B- and T-cell-mediated responses elicited during primary SARS-CoV-2 infection could reduce susceptibility to and infectiousness of the following infections and offer protection against severe disease, i.e. COVID-19 [64]. The estimation of the model parameters and evaluation of relaxation strategies in light of waning of sterilizing immunity lies outside the scope of our study but it should be addressed in future work when convincing data on reinfections in unvaccinated and vaccinated individuals become available.

Another limitation is that our results are based on early data on the efficacy in clinical trials and real-world effectiveness of the Pfizer-BioNTech vaccine [15, 18–22]. We also assume that vaccine efficacy against the B.1.1.7 variant circulating in Portugal is the same as the efficacy reported from studies conducted in other locations as supported by the recent study among working age adults in England [22], where the dominant variant in circulation was B.1.1.7. This study demonstrated that effectiveness of the Pfizer-BioNTech vaccine against symptomatic and asymptomatic infection is 86% seven days after two doses [22]. However, SARS-CoV-2 mass vaccination programmes and prolonged control measures can generate selection pressure leading to viral adaptation, antigenic divergence or vaccine escape. Viral adaptations may contribute to decreasing efficacy of existing vaccines via faster waning of sterilizing immunity. For example, recent experiments demonstrate that the South African variant B.1.351 shows reduced neutralizing antibody binding increasing the prospects of reinfection and hampering the efficacy of spike-based vaccines [65]. This will need consideration in vaccine development and evaluation of future vaccination programmes and relaxation scenarios in mathematical transmission models. A possible case where an antigenic escape variant caused a resurgence of COVID-19 despite high population-level seroprevalence was observed in Manaus, Brazil [30].

To summarize, our study provides timely input into the discussion about the pandemic response during the vaccination rollout in Portugal. Our analyses suggest that the pressing need to restart socioeconomic activities might lead to new waves of hospitalizations in 2021 and that substantial measures prove necessary to control COVID-19 throughout 2021. More favourable scenarios that help to avoid future waves include relaxation of measures as in summer 2020 or a step-wise approach when measures are relaxed gradually until the end of 2021.

## Methods

### Overview

The transmission model was calibrated using a combination of behavioral, surveillance and demographic data for Portugal. Parameter estimates were obtained from the model fit to (i) age-stratified COVID-19 hospitalization data (*n* = 28, 482) in the period from 26 February 2020 till 15 January 2021 and (ii) cross-sectional age-stratified SARS-CoV-2 seroprevalence data (*n* = 2, 301) assessed from 21 May 2020 till 8 July 2020 [54]. The model was further used to investigate relaxation scenarios as vaccination is rolled out in 2021.

### Data

The hospitalization data included *n* = 28, 482 COVID-19 hospitalizations longer than 24 hours by date of admission and stratified by age during the period of 325 days following the first official case in Portugal (2 March 2020). The data was padded with 5 days without hospitalizations (from 26 February till 1 March 2020) to allow for the estimation of the number of infected individuals at the start of the pandemic. The hospitalization data spanned the first wave in spring 2020, relatively low epidemic activity in summer 2020, the second wave that started in autumn 2020 till mid-December 2020 and the third wave that started in mid-December 2020 and was still ongoing on 15 January 2021. The data source for hospital data was the Central Administration of the Health System and the Shared Services of the Ministry of Health, covering all public hospitals in Portugal receiving COVID-19 patients. Since early in the pandemic, Portugal adopted a policy of hospitalizing only patients who did not gather minimum conditions for being followed at the domicile, either due to clinical or sanitary conditions. This policy has not changed during the course of the pandemic.

The SARS-CoV-2 seroprevalence data was based on the First National Serological Survey (ISNCOVID-19) in Portugal in May/July 2020 [54]. This cross-sectional seroepidemiological survey was conducted on a sample of *n* = 2, 301 Portuguese residents, aged 1 year or older, after the first wave. The survey sample was selected using a two-stage stratified non-probability sampling design (quota sampling) [54]. SARS-CoV-2 IgM and IgG antibodies were measured in serum samples by enzyme-linked immunosorbent assay. Further details of the study are given in [54]. For the model fitting, we used the sample size, the number of positive samples and 95% confidence intervals stratified by age group reported in [54].

The demographic composition of the Portuguese residents was taken for 2019 from the Contemporary Portugal Database (Pordata) [66]. The vaccination analyses made use of the vaccination programme (Table 1), as defined by the Directorate-General of Health [51]. The programme defines vaccine uptake prioritization by age and morbidities and runs in three phases from 27 December 2020 till 31 December 2021. The age distribution of morbidities in the Portuguese population was extracted from the Shared Services of the Ministry of Health on the basis of ICPC-2 (International Classification of Primary Care) codes (Table S1). The vaccination rollout data for Portugal was taken from the ECDC website.

The baseline (pre-pandemic) contact matrices for transmission-relevant contacts for Portugal were taken from the recent study by Mistry and colleagues [67]. The contact matrix for Portugal after the introduction of measures to control the first wave of hospitalizations (April 2020) was inferred using the contact matrix for the Netherlands based on a cross-sectional survey carried out in April 2020 (PIENTER Corona study) [68].

### Transmission model

We extended an age-stratified SARS-CoV-2 transmission model from [43] to include vaccination (Figure 8). The model has susceptible-exposed-infectious-recovered structure, whereby susceptible persons (*S*) may become latently infected (*E*) before progressing to become infectious (*I*). Infectious persons either get hospitalized (*H*) or recover without hospitalization (*R*). Disease-related mortality and discharge from the hospital are not explicitly modeled. Therefore, the *H*-compartment contains the cumulative number of persons who experience severe symptoms and recover (or die) after admission to the hospital. Similarly, the *R*-compartment contains the cumulative number of persons who recover after having mild or no symptoms. The force of infection is given by a weighted sum of the fraction of the infectious population in different age groups (red dashed boxes in Figure 8). We consider a stable population and thus do not include natural birth and death processes. The contact rates, forces of infection, susceptibilities and hospitalization rates are age-specific.

**Figure 8.**
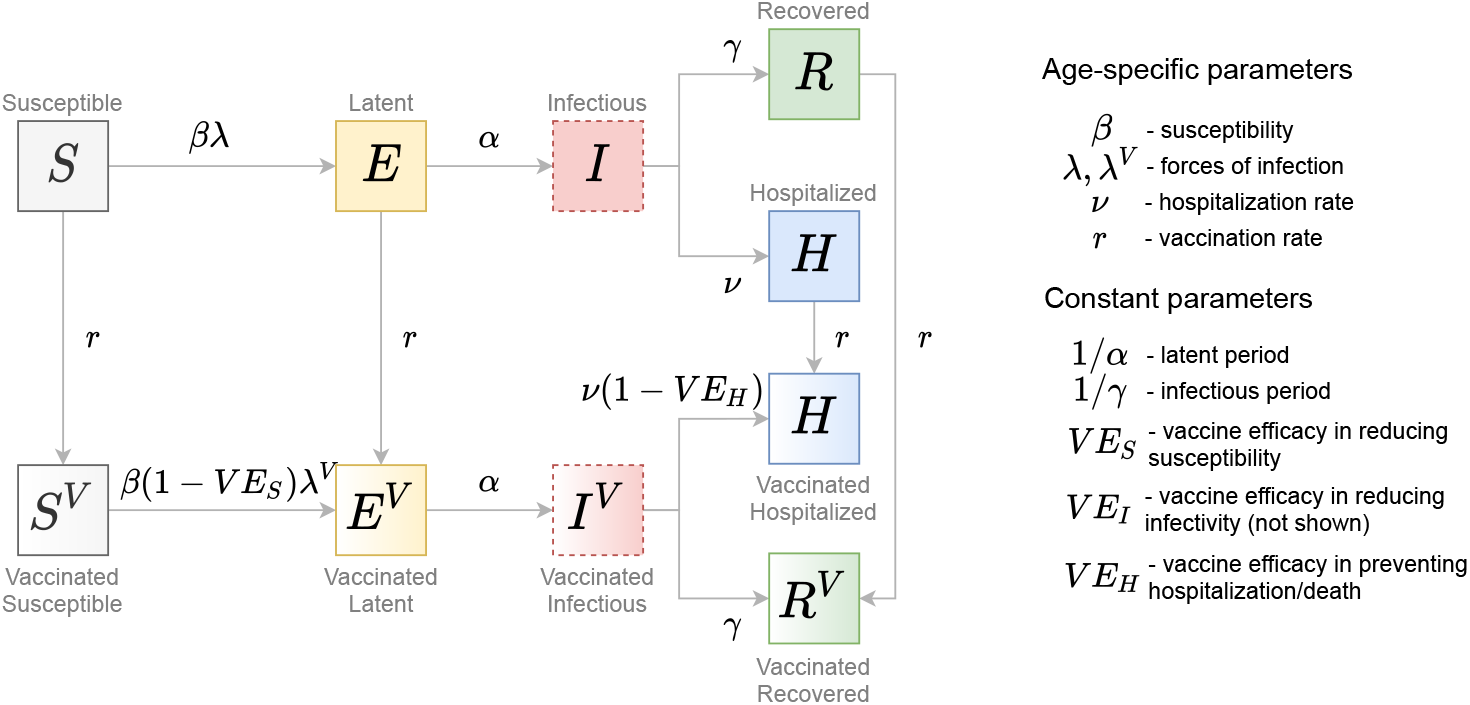
Schematic of the transmission model. Gray arrows show epidemiological transitions. Red dashed boxes indicate compartments contributing to the forces of infection. The model is age-structured and involves an extended SEIR-type framework. Vaccinated persons may experience behavior compensation post-vaccination modelled as a return to pre-pandemic contact rates among vaccinated persons as compared to unvaccinated persons who may continue to have reduced contact rates due to control measures. The vaccine has three effects: (i) reduction in susceptibility of vaccinated relative to unvaccinated (*V E*_*S*_); (ii) reduction in infectivity of vaccinated relative to unvaccinated (*V E*_*I*_, not shown); (iii) reduction in hospitalization rate of vaccinated relative to unvaccinated (*V E*_*H*_).

In line with the current guidelines, we assume that vaccine can be delivered to all people independently from their disease history with the exception of those who might be currently infectious (*I*-compartment). Not vaccinating infectious compartment implies that vaccine is not given to asymptomatic persons but these represent a small fraction of the population at any given time. We also vaccinate the *H*-compartment as this compartment comprises everyone who has ever been admitted to hospital. Whilst this assumption means that the currently hospitalized persons are vaccinated too, their number is very small compared to the total number of people in the *H*-compartment. The vaccine has three mechanisms of action: (i) reducing susceptibility (*V E*_*S*_); (ii) reducing infectivity (*V E*_*I*_); (iii) reducing hospitalization rate (*V E*_*H*_). The vaccine has no effect in persons who recovered from natural infection (*R* and *H* compartments). We assume that protection after vaccination is achieved immediately and is equivalent to two vaccine doses, and that the duration of protection after both natural infection and vaccination is about two years (time horizon of our analyses). Finally, we allow for behavior compensation post-vaccination modelled as a return to pre-pandemic contact rates among vaccinated persons as compared to unvaccinated persons who may continue to have reduced contact rates due to control measures. This is reflected in generally different forces of infection for unvaccinated and vaccinated persons. The full description of the model parameters is given in Tables S2 and S4.

### Model equations

The model was implemented in Mathematica 10.0.2.0 using a system of ordinary differential equations for the number of persons in different compartments shown in Figure 1. The transmission model was stratified into *n* = 10 age groups: [0, 5), [5, 10), [10, 20), [20, 30), [30, 40), [40, 50), [50, 60), [60, 70), [70, 80), 80+.

The equations for the numbers of unvaccinated persons in age group *k, k* = 1, …, *n*, who are susceptible (*S*_*k*_), exposed (*E*_*k*_), infectious (*I*_*k*_), recovered (*R*_*k*_) and hospitalized (*H*_*k*_) read as follows

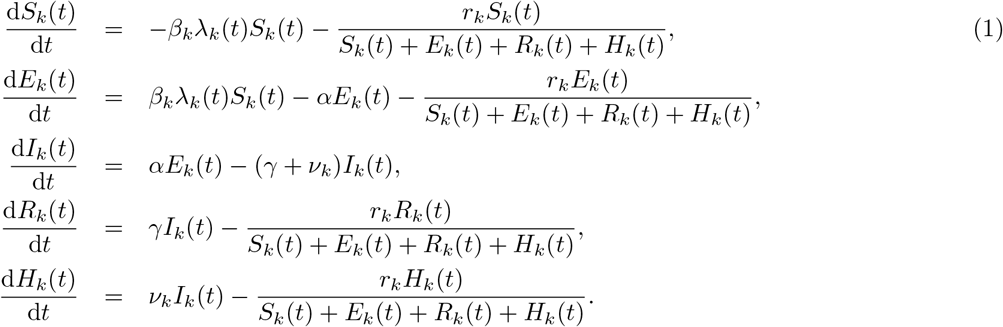

The equations for the numbers of vaccinated persons in age group *k* who are vaccinated susceptible 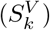, exposed 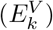, infectious 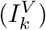, recovered 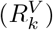 and hospitalized 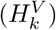 are given by

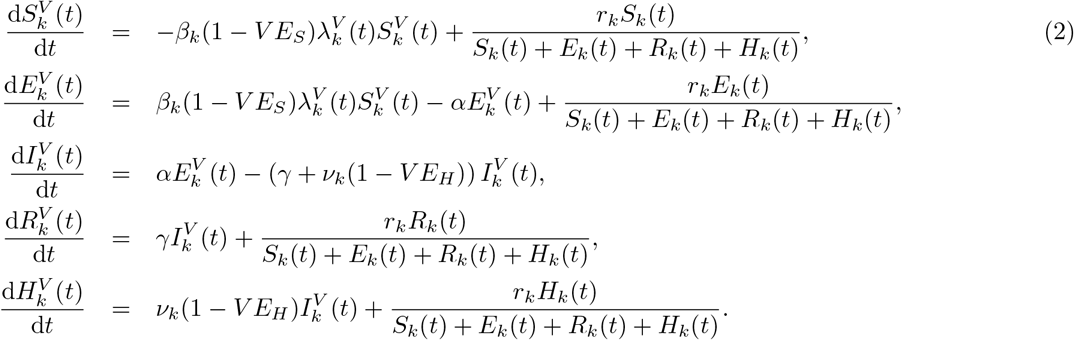

Persons get vaccinated in *S, E, R* and *H* states. The vaccination rates *r*_*k*_ are age-specific. We denote the contact rate of an unvaccinated person in age group *k* with persons in age group *l, c*_*kl*_(*t*), and the contact rate of a vaccinated person in age group *k* with persons in age group *l*, 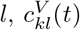. The forces of infection for unvaccinated and vaccinated persons are given by

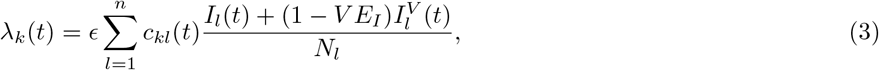

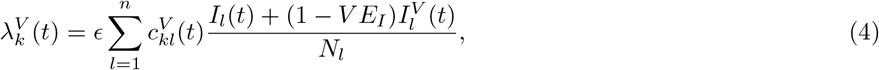

where *N*_*k*_ is the number of individuals in age group *k*, 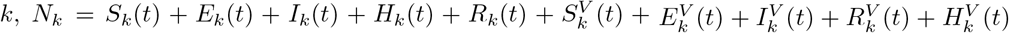. Note that Eqs. (3) and (4) imply that the entire population participates in the contact process including persons in the *H*-compartment but that *H*-persons are not infectious. This is based on the fact that the vast majority of people in the *H*-compartment are recovered after hospitalization, and a very small proportion is currently hospitalized. We assume that currently hospitalized persons continue to have contacts with the personnel and visitors but they cannot infect them because of the use of individual protective measures.

The initial condition for the model was 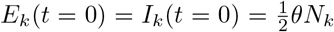 and *S*_*k*_(*t* = 0) = (1 - *θ*)*N*_*k*_, where *t* = 0 is 26 February 2020. The parameter *θ* denotes the initial fraction of the population that was infected (split equally between infectious and exposed). This parameter accounts for importation of new cases at the start of the pandemic and was estimated jointly with other parameters. Importation of cases was not implemented at later stages of the pandemic due to a large pool of infectious individuals within the country.

The rapid spread of B.1.1.7 variant, that is estimated to be about 50% more transmissible based on the data from England [5–7], fueled the third wave of hospitalizations in Portugal. The increasing dominance of this variant was modelled empirically as a gradual increase in the probably of transmission per contact by 50% as follows 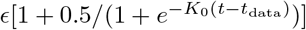, where *E* and *K*_0_ were estimated based on the data until 15 January 2021 (Figure S2) and *t*_data_ is the last date in the hospital admission data (15 January 2021).

### Observation model and parameter estimation

To generate a set of plausible parameters and initial conditions for our projections, we fitted the model to hospitalization data and serological testing data, using a similar approach as before [43, 69]. We incorporated the transmission model, Eq. (1), in a Bayesian statistical model with likelihood function constructed as follows. Let *h*_*k,m*_ denote the observed number of hospitalizations in age group *k* and day *t*_*m*_. The expected number of hospitalizations during day *t*_*m*_ is approximately equal to 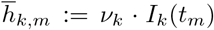. To account for reporting errors and heterogeneity in the hospitalization rate within age groups, we assume that *h*_*k,m*_ has a negative-binomial distribution with mean 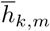 and variance 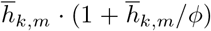. The parameter *φ* determines the overdispersion of the reporting of hospitalizations. The hospitalization data were stratified into the ten age groups [0, 5), [5, 10), [10, 20), [20, 30), [30, 40), [40, 50), [50, 60), [60, 70), [70, 80), 80+.

The seroprevalence data were stratified into the five age groups [1, 10), [10, 20), [20, 40), [40, 60) and 60+ [54]. Hence, for the hospitalization data and the transmission model, a finer age stratification is used than for the seroprevalence data. We assume that individuals in seroprevalence age group 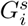 were sampled from hospitalization age class 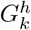 with probability *p*_*ik*_ proportional to the relative population size of 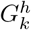 compared to 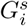.

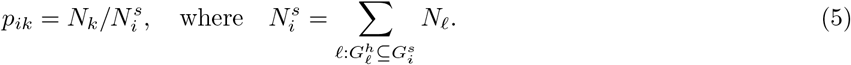

As before [43], we assume that the seroprevalence data represents a random sample from each age group. Hence, the number of positive samples *f*_*i*_ has a binomial distribution with population size *L*_*i*_, equal to the total number of samples for age class *i*, and success probability *q*_*i*_. The success probability is defined in terms of the fraction of susceptible individuals *S*_*k*_(*T*) at sampling time *T* and the probabilities *p*_*ik*_:

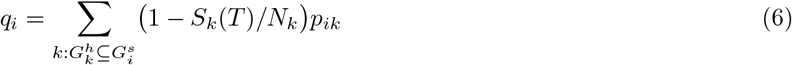

To account for the fact that no children below the age of 1 year were included in the serology samples, we reduced the population size *N*_1_ with the size of the age group [0, 1) (86, 579 persons) in Eq. (6) and Eq. (5).

The prior distribution of the model is specified in Table S3. The model was fitted with Stan [70] in R 3.6.0 and R Studio 1.3.1056. We used 4 parallel chains, each of length 1,000, with a warm-up period of 500, resulting in 2,000 samples from the posterior distribution. Convergence was assessed with the Gelman-Rubin 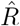-statistic, which was close to 1 for all parameters. The estimated model parameters are shown in Figures S1 and S2.

### Time-varying contact patterns

The contact patterns in the population varied with time due to introduction/reinforcement or relaxation of control measures as follows: 0) introduction of measures to control the first pandemic wave (first lockdown, March 2020); 1) relaxation of measures after the first wave was curbed (May 2020); 2) further relaxation of measures that included school opening (September 2020); 3) reinforcement of measures to control the second wave (second lockdown, November 2020); 4) relaxation of measures around Christmas 2020; 5) reinforcement of measures to control the third wave (third lockdown, January 2021).

We denote *c*_*kl*_(*t*) the contact rate for a person in age group *k* (*k* = 1, …, *n*) with persons in age group *l* (*l* = 1, …, *n*) at time *t*. The contact rate denotes the number of transmission-relevant contacts per day such as touching or having a conversation with someone [67, 68]. Our fitting procedure allows to estimate *c*_*kl*_(*t*) by assuming that changes due to control measures described in 0)-5) occur as a series of smooth transitions.

To describe the transition 0) from the baseline (pre-pandemic) contact rate *b*_*kl*_ to the contact rate after the first lockdown *a*_*kl*_ we write down *c*_*kl*_(*t*) as a linear combination of contact rates *b*_*kl*_ and *a*_*kl*_ with coefficients constructed using a logistic function 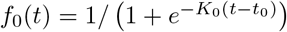 as follows

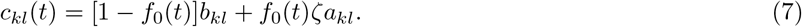

The parameter *K*_0_ of the logistic function describes the speed with which the first lockdown is enforced. The parameter *t*_0_ describes the mid-time of the introduction of the first lockdown. Note in Eq. 7 we introduced the factor *ζ* ∈ [0, 1] to reflect that not all reported contacts after the first lockdown might be relevant for transmission, for example, due to mask-wearing or physical distancing when a contact took place. Therefore, the baseline (prepandemic) contact rates are described by the matrix *b*_*kl*_, and the contact rates after the first lockdown are described by the matrix *ζa*_*kl*_.

The pre-pandemic matrix *b*_*kl*_ for Portugal was taken from [67] (Figure 9 **a**). The matrix after the first lockdown *a*_*kl*_ was inferred using the contact matrix for the Netherlands based on a cross-sectional survey carried out in April 2020 (PIENTER Corona study) [68]. Since measures enforced during the first lockdown in the two countries were similar (e.g., all schools were closed, all non-essential work was done from home etc.) we reduced the age-specific

**Figure 9.**
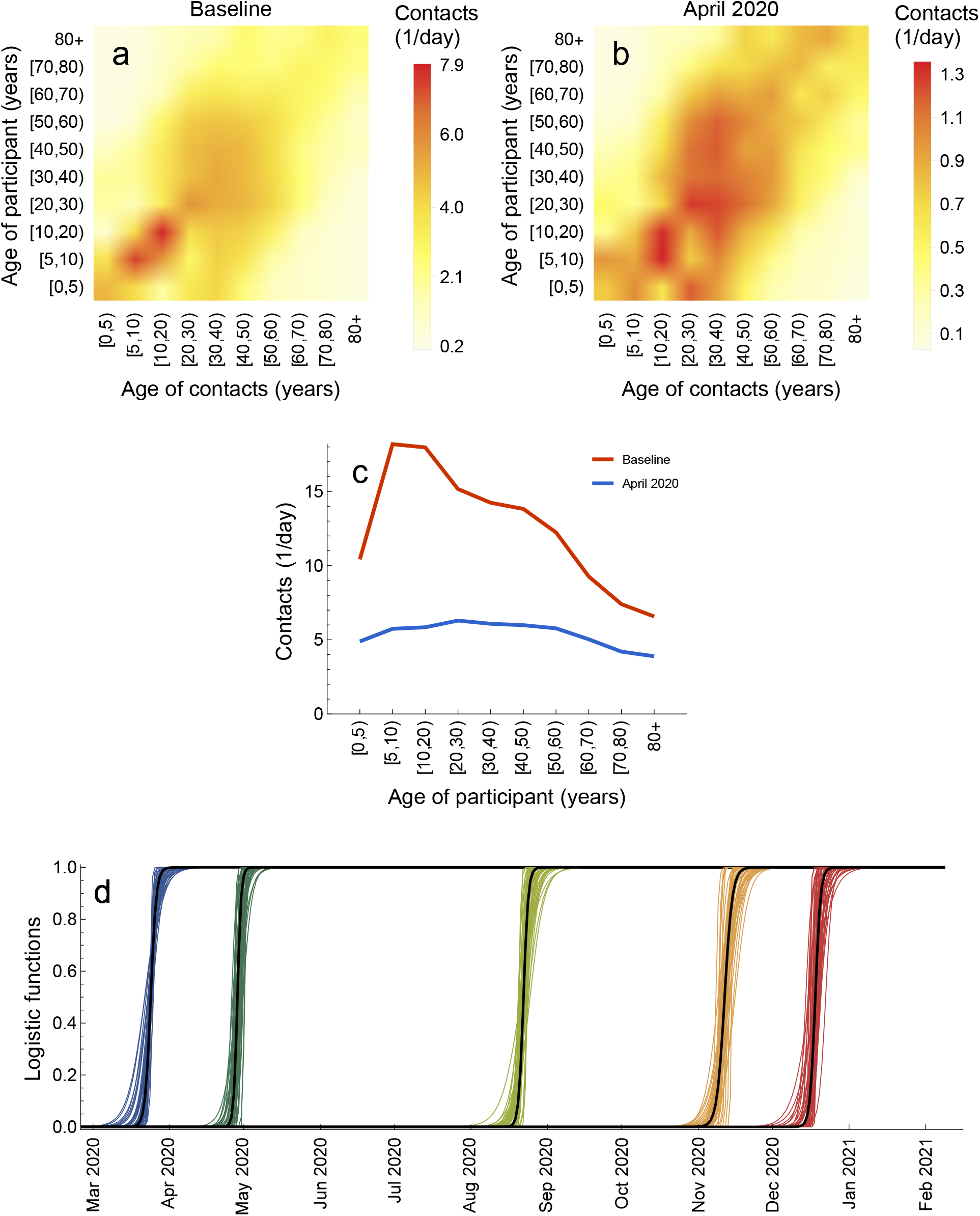
Contact matrices. **a** Baseline (pre-pandemic) contact matrix. **b** Contact matrix after the introduction of measures in April 2020. **c** Average number of contacts for a person in a given age group. **d** Logistic functions describing transitions between contact matrices. Shown are *f*_0_ (blue), *f*_1_ (dark green), *f*_2_ (light green), *f*_3_ (orange), and *f*_4_ (red) based on 50 samples from the posterior distribution. contact rates for Portugal after the lockdown by the same percentage as it was observed in the Netherlands (Figure 9 **b**). The resulting number of daily contacts for a person in given age group at baseline and after the lockdown in April 2020 is shown in Figure 9 **c**. Like for the Netherlands [68], we observe larger reductions in contacts for children (due to school closure) and smaller reductions for elderly because most of their contacts were essential (e.g., with healthcare personnel or caretakers) and thus were not affected by the lockdown. The parameter *ζ* that multiplies the inferred matrix *a*_*kl*_ can account for discrepancies between the real and inferred matrix.

To describe the contact rates after transitions 1)-4) have taken place, we assume that these can be written as a liner combination *u*_*i*_*b*_*kl*_ + (1 - *u*_*i*_)*ζa*_*kl*_, *i* = 1, …, 4, where *u*_*i*_ is the proportion of time a person behaves as before the pandemic and (1 - *u*_*i*_) is, respectively, the proportion of time a person behaves as during the first lockdown. This contact structure can, therefore, interpolate between the first (most strict) lockdown and no measures in place at all. Since the third lockdown was similar to the first lockdown, the transition 5) was modelled as a return to the lockdown contact matrix *ζb*_*kl*_. As before, the transitions between the contact rates during periods 1)-5) are modelled using logistic functions 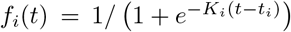, where *i* = 1, …, 5. The general contact rate can therefore be written as

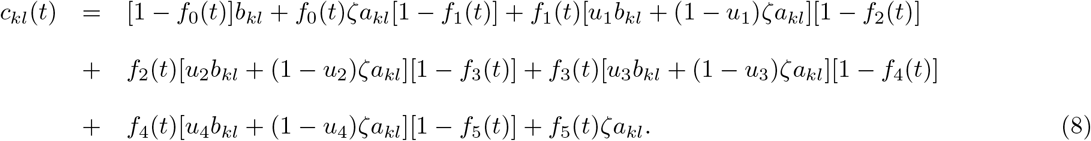

All the parameters that describe *c*_*kl*_(*t*), except for the last transition 5) for which hospitalization data are not available, are estimated (Table S4). The estimates for these 15 parameters *ζ, u*_*i*_ (*i* = 1, …, 4), *t*_*i*_ (*i* = 0, …, 4) and *K*_*i*_ (*i* = 0, …, 4) are shown in Figure S2. The estimated logistic functions are plotted in Figure 9 **d**.

In the main analyses (Figures 6 and 7), the contact rates for vaccinated persons were equal to those unvaccinated, 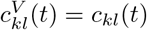. In the sensitivity analyses (Figure S7), they were set to pre-pandemic contacts as follows, 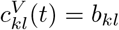. The contact rate presented in Figures 3, 6 and 7 was the average contact rate in the population calculated as follows 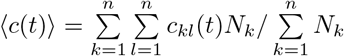. Note that this expression makes use of the fact that in the main analyses 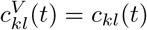.

The relaxation scenarios during the vaccination rollout are modelled as a transition from the contact rate described by Eq. (8) to the contact rate *b*_*kl*_ (Scenario 1); *u*_2_*b*_*kl*_ + (1 - *u*_2_)*ζa*_*kl*_ (Scenario 2); *u*_1_*b*_*kl*_ + (1 - *u*_1_)*ζa*_*kl*_ (Scenario 3); *u*_1_*b*_*kl*_ + (1 - *u*_1_)*ζa*_*kl*_ (Scenario 4, Step 1); *u*_2_*b*_*kl*_ + (1 - *u*_2_)*ζa*_*kl*_ (Scenario 4, Step 2); *b*_*kl*_ (Scenario 4, Step 3). The parameters of the logistic functions describing these transitions are specified in Table S4.

### Time-varying effective reproduction number

The basic reproduction number, *R*_0_, is the average number of secondary infections caused by a single infectious individual at the beginning of the epidemic in a disease-free, totally susceptible population. If *R*_0_ *>* 1 the disease will spread exponentially. If *R*_0_ < 1 the number of infectious persons declines exponentially and the disease is not able to spread. In general, *R*_0_ depends on the type of virus but also on the contact patterns in the population.

When the disease has already spread and we have no longer a fully susceptible population but some part of the population is immune due to natural infection or vaccination, the generalization of *R*_0_ is given by the effective reproduction number, *R*_*e*_(*t*). *R*_*e*_(*t*) depends on the type of virus, the level of population immunity and the contact patterns in the population. The full control of the disease is achieved when *R*_*e*_(*t*) < 1 and the contact rates in the population are at their pre-pandemic levels, i.e., not anymore affected by control measures. A partial control is achieved when *R*_*e*_(*t*) < 1 but the contact rates have not been restored to their pre-pandemic levels yet as is currently the case for SARS-CoV-2 in Portugal.

In a deterministic compartmental model such as the one employed here, the calculation of *R*_0_ and *R*_*e*_(*t*) can be performed using the next-generation matrix (NGM) method [71]. The starting point of the method is to calculate the Jacobian **J** of the equations for the latent 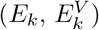 and infectious 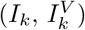 age classes *k, k* = 1, …, *n*, isolated from the full model given by Eqs. (1) and (2). The Jacobian **J** is then evaluated at the disease-free equilibrium of interest.

For *R*_0_ calculation, the disease-free equilibrium is

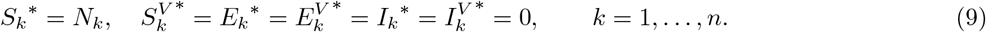

For *R*_*e*_(*t*) calculation with or without vaccination, the disease-free equilibrium is

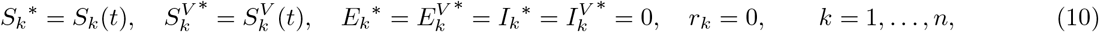

where the time-dependent variables *S*_*k*_(*t*) and 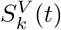 are obtained from the solutions of the full model given by Eqs. (1) and (2).

Following [71], the Jacobian **J** may be recast as follows

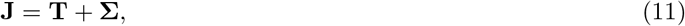

where the transmissions matrix **T** contains the terms associated with the production of new infections, and the transitions matrix **Σ** contains the terms associated with all other state changes. After performing this operation, we construct a new matrix **K**_**L**_, called the large domain NGM [71], given by

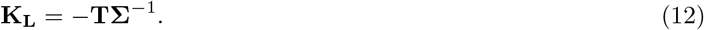

The basic reproduction number *R*_0_ at time *t* = 0 and the effective reproduction number *R*_*e*_(*t*) at any time *t* are given by the spectral radius of **K**_**L**_ which is the largest eigenvalue of **K**_**L**_. For the purpose of computing the spectral radius, **K**_**L**_ can be further reduced as detailed in [71]. The explicit expressions for matrices **J, T, Σ** and **K**_**L**_ are given in the Mathematica notebooks available in the GitHub repository, https://github.com/lynxgav/COVID19-vaccination.

### Population immunity

The unprotected population was computed as the number of individuals in the fully susceptible compartment *S* (Figure 8). The population protected by natural infection was computed as all individuals arriving into the infectious compartment *I*, independently of whether these individuals will or will not be vaccinated later on. Recall, that in the model vaccine has no effect in individuals who are recovered from natural infection and, therefore, the population protected by vaccination grows slower than vaccination coverage. The population protected by vaccination was computed as all individuals arriving into the compartments *S*^*V*^ and *I*^*V*^ due to vaccination.

### Vaccine efficacies

Vaccine efficacies in reducing susceptibility (*V E*_*S*_), infectivity (*V E*_*I*_) and hospitalization rate (*V E*_*H*_) were set using initial data from clinical trials and real-word studies for the Pfizer-BioNTech vaccine [15, 18–22]. Important to note, that the efficacies reported in all these studies are not conditioned on infection while they are in the models like ours. For a more complete discussion on this topic, we refer the reader to the pedagogical work by Lipsitch and Kahn [23] and the report for England by the Scientific Advisory Group for Emergencies [26].

The vaccine efficacy in reducing susceptibility (*V E*_*S*_) was set based on vaccine efficacies and effectiveness against infection (*V E*_infection_) reported in clinical trials and real-word studies, i.e.

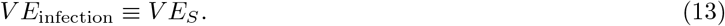

The vaccine efficacy in reducing infectivity (*V E*_*I*_) was assumed to be the same as vaccine efficacy in reducing disease conditioned on infection (*V E*_disease|infection_), i.e. *V E*_disease|infection_ ≡*V E*_*I*_. *V E*_disease|infection_ was calculated using the efficacy against disease (*V E*_disease_) reported in clinical trials as follows

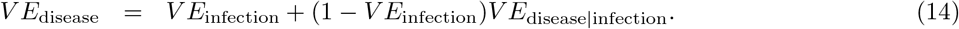

The vaccine efficacy in reducing hospitalization rate (*V E*_*H*_) is equal to vaccine efficacy against severe disease conditioned on disease (*V E*_severe disease|disease_), i.e. *V E*_severe disease|disease_ ≡*V EH*. *V E*_severe disease|disease_ was calculated using the vaccine efficacy against severe disease (*V E*_severe disease_) reported in trials as follows

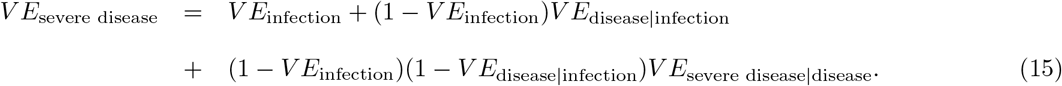

We used an optimistic and a pessimistic set of vaccine efficacies for *V E*_*S*_, *V E*_*I*_ and *V E*_*H*_ (Table S2) based on the range of values for *V E*_infection_, *V E*_disease_, and *V E*_severe disease_ reported in the literature [15, 18–22]. For the optimistic set explored in the main analyses (Figures 6 and 7), we used *V E*_infection_ = 94%, *V E*_disease_ = 94%, and *V E*_severe disease_ = 98% (corresponding to *V E*_*S*_ = 94%, *V E*_*I*_ = 0%, and *V E*_*H*_ = 67%) [15, 18, 19, 21, 22, 26]. For the pessimistic set explored in sensitivity analyses (Figure S7), we used *V E*_infection_ = 55%, *V E*_disease_ = 55%, and *V E*_severe disease_ = 55% (corresponding to *V E*_*S*_ = 55%, *V E*_*I*_ = 0%, and *V E*_*H*_ = 0%) [19, 20, 26]. Other efficacies reported in the literature for the Pfizer-BioNTech vaccine and other existing vaccines fall in between the optimistic and pessimistic values we used. This broad range of values is also relevant in case the market share of different vaccine brands in Portugal gets changed throughout 2021.

## Data Availability

The data used in this study are publicly available at https://github.com/lynxgav/COVID19-vaccination.

## Code availability

The codes reproducing the results of this study are publicly available at https://github.com/lynxgav/COVID19-vaccination.

## Acknowledgements

G.R., J.V., A.N., M.C.G. were supported by FCT project reference 131 596787873, awarded to G.R. M.V. was supported by the European Union H2020 ERA project (No 667824 EXCELLtoINNOV). The contribution of C.H.v.D. was under the auspices of the US Department of Energy (contract number 89233218CNA000001) and supported by the National Institutes of Health (grant number R01-OD011095).

## Author contributions

G.R. conceived and supervised the study. G.R. and J.V. developed the transmission model. C.H.v.D. developed the observation model. J.V. conducted preliminary model analyses. G.R. conducted all final analyses, prepared figures and wrote the manuscript. A.N., M.C.G., M.v.B., M.E.K, and M.V. provided data, validated the model and analyses. All authors contributed to interpretation of the results, writing the final version of the manuscript and gave final approval for publication.

## Additional information

Supplementary Figures and Tables are given at the end of the manuscript.

## Supplementary Figures and Tables

**Figure S1.**
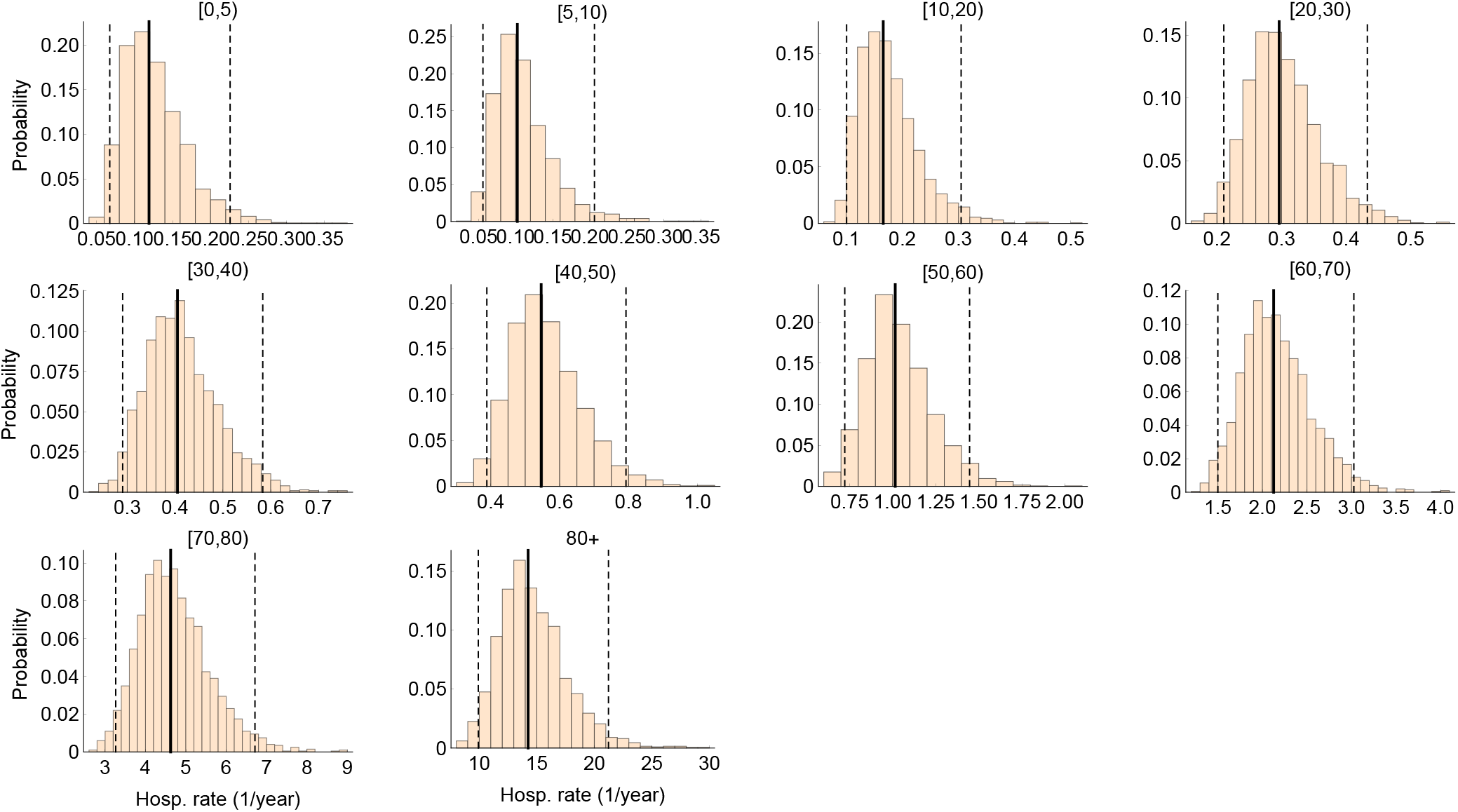
Estimated hospitalization rates. The histograms of age-specific hospitalization rates estimated by the model. The solid and the dashed lines are, respectively, the medians and the 95% credible intervals based on 2,000 parameter samples from the posterior distribution.

**Figure S2.**
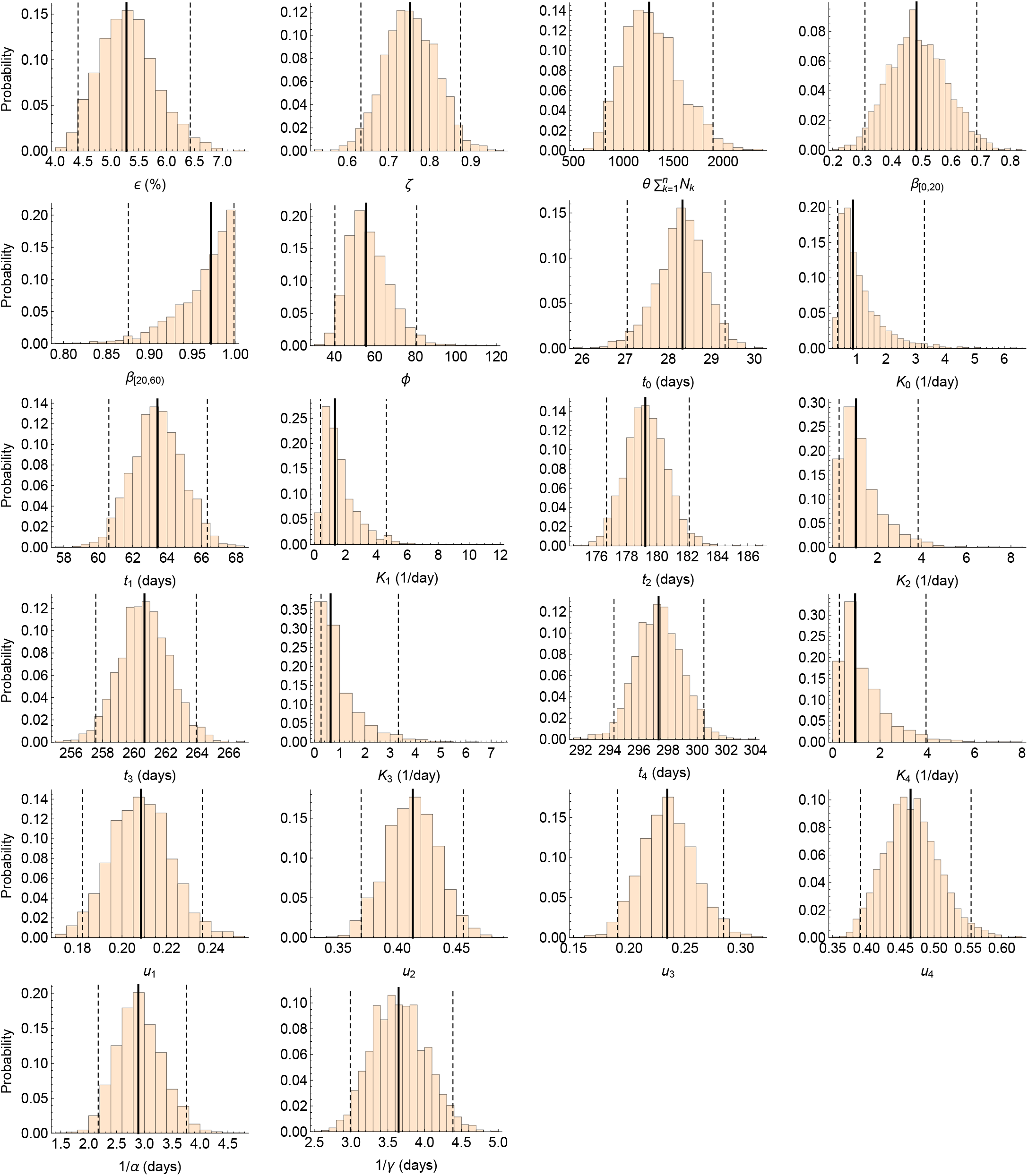
Estimated model parameters. The histograms of model parameter estimates. The solid and the dashed lines are, respectively, the medians and the 95% credible intervals based on 2,000 parameter samples from the posterior distribution. Time *t* = 0 corresponds to 26 February 2020.

**Figure S3.**
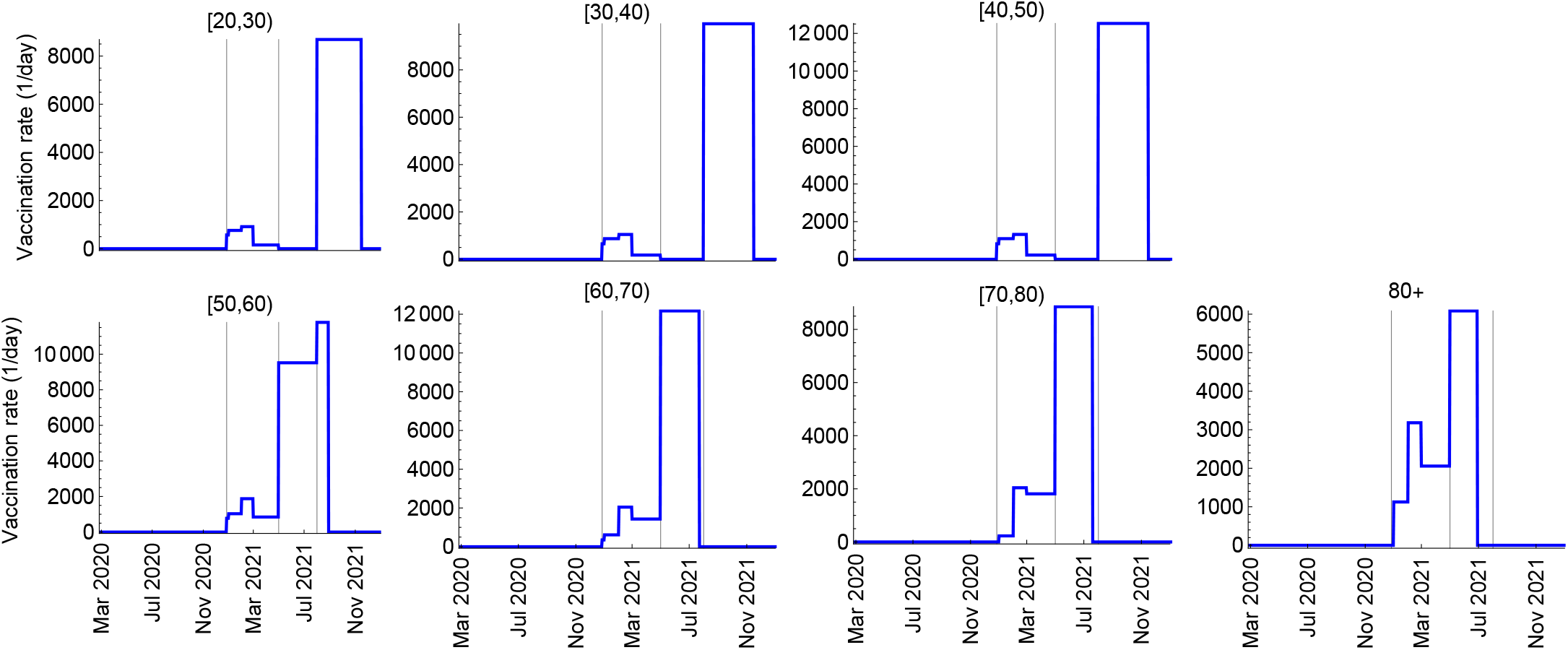
Age-specific vaccination rates. Vaccination rate (number of persons vaccinated per day) per age group calculated using the national vaccination plan (Table 1) and age distribution of various vaccination categories (Figure 4 **a**). The vertical lines indicate the starting dates of different phases of vaccination (Table 1). According to the current guidelines persons under 18 years old are not eligible for vaccination. In the model, we assumed that the age group of 0 to 20 years old is not vaccinated.

**Figure S4.**
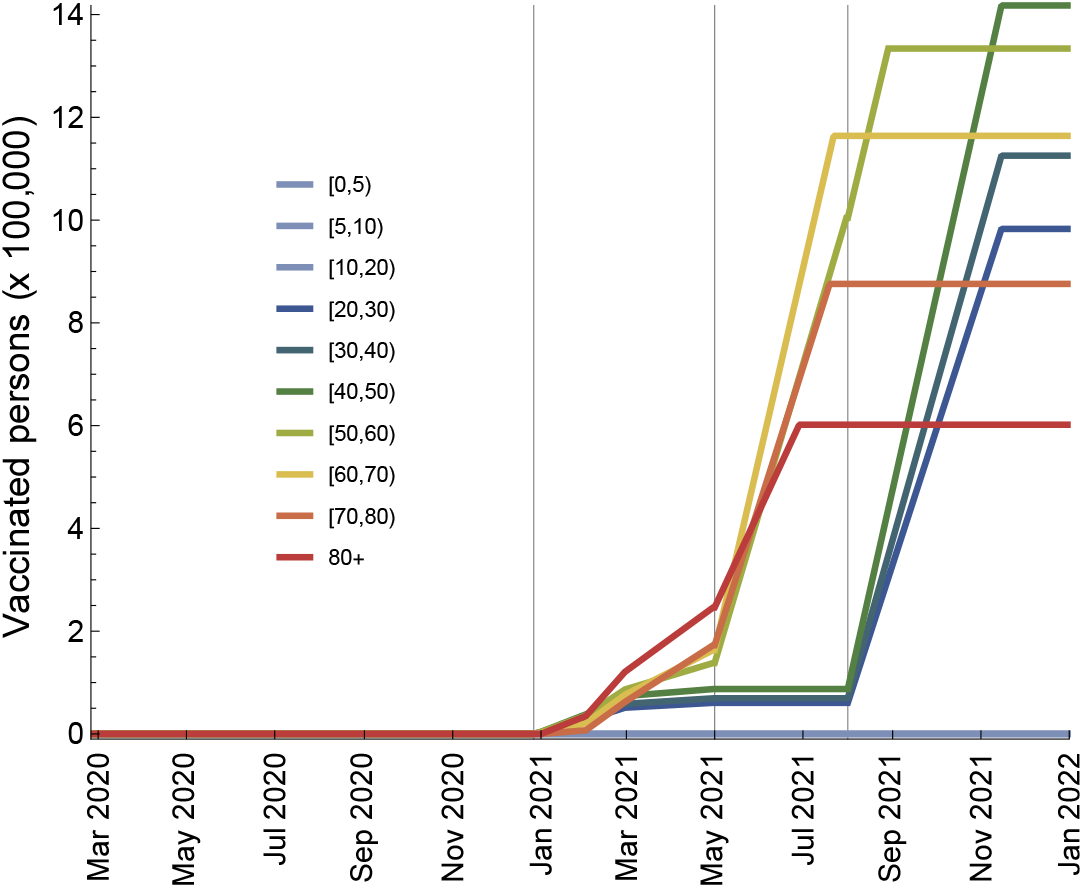
Number of vaccinated persons per age group during the vaccination rollout. These numbers were calculated using the national vaccination plan (Table 1) and age distribution of various vaccination categories (Figure 4 **a**). The vertical lines indicate the starting dates for vaccination of different phases of vaccination (Table 1). According to the current guidelines persons under 18 years old are not eligible for vaccination. In the model, we assumed that the age group of 0 to 20 years old is not vaccinated.

**Figure S5.**
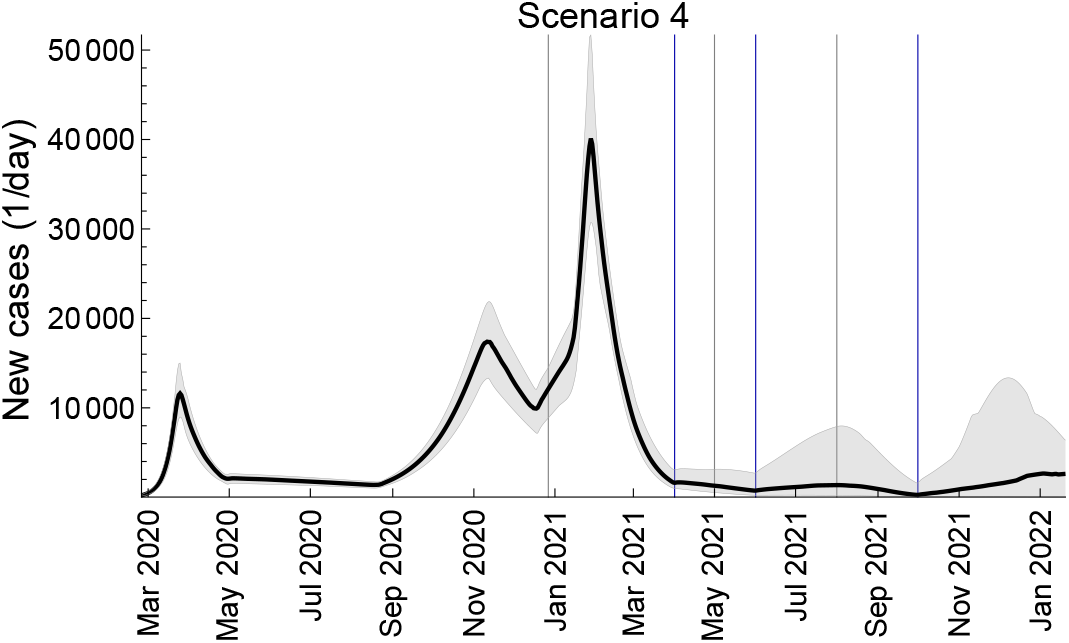
Infectious cases dynamics. New daily cases of SARS-CoV-2 for Scenario 4 presented in Figure 7 in the main text. The black line is the median trajectory estimated from the model. The gray shaded region corresponds to 95% credible intervals. The blue vertical lines indicate the mid-points of relaxation steps (1 April, 1 June, 1 October). The gray vertical lines indicate the starting dates for different vaccination phases (Table 1).

**Figure S6.**
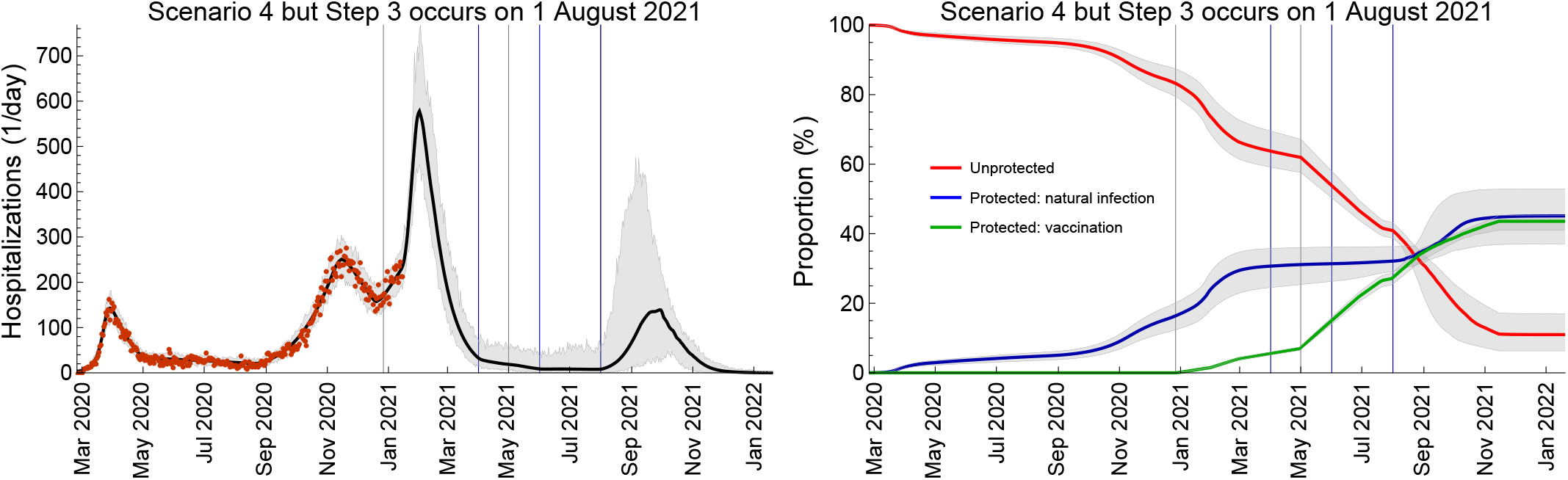
Impact of timings of different relaxation steps. Total daily hospital admissions with COVID-19 and proportion of protected population for Scenario 4 (Figure 7 in the main text) with Step 3 occurring on 1 August instead of 1 October 2021. The hospitalization data are shown as red dots. The solid lines are the median trajectories estimated from the model. The gray shaded regions correspond to 95% credible intervals. The blue vertical lines indicate the mid-points of relaxation steps (1 April, 1 June, 1 August). The gray vertical lines indicate the starting dates for different vaccination phases (Table 1).

**Figure S7.**
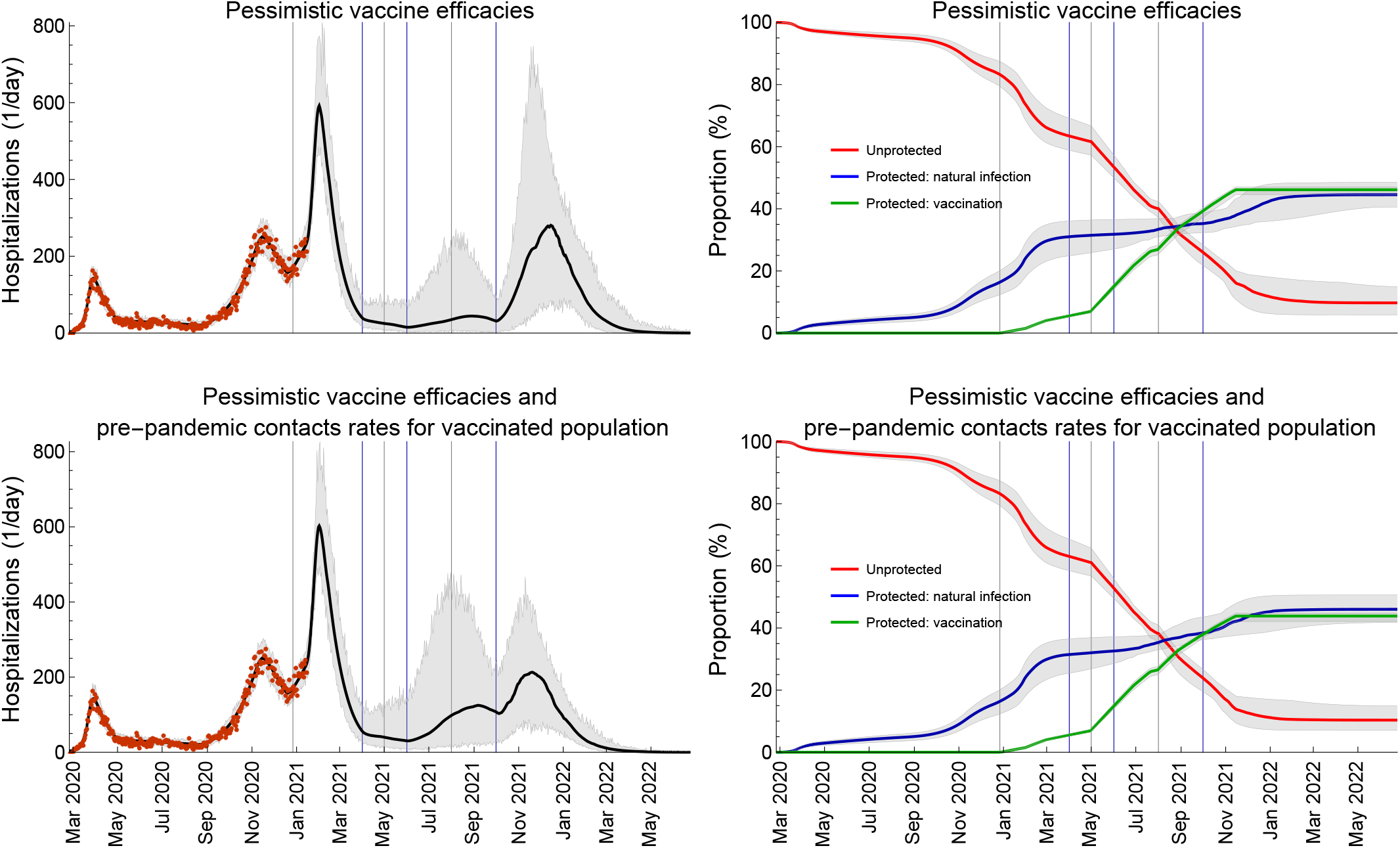
Impact of vaccine efficacies and contact rates of vaccinated individuals. Scenario 4 (Figure 7 in the main text) but with a pessimistic set of vaccine efficacies (Table S2). In addition to using a pessimistic set of vaccine efficacies, we allow for behavior compensation post-vaccination modelled as a return to pre-pandemic contact rates among vaccinated persons as compared to unvaccinated persons who may continue to have reduced contact rates due to control measures. The hospitalization data are shown as red dots. The solid lines are the median trajectories estimated from the model. The gray shaded regions correspond to 95% credible intervals. The blue vertical lines indicate the mid-points of relaxation steps (1 April, 1 June, 1 October 2021). The gray vertical lines indicate the starting dates for different vaccination phases (Table 1).

**Table S1.**
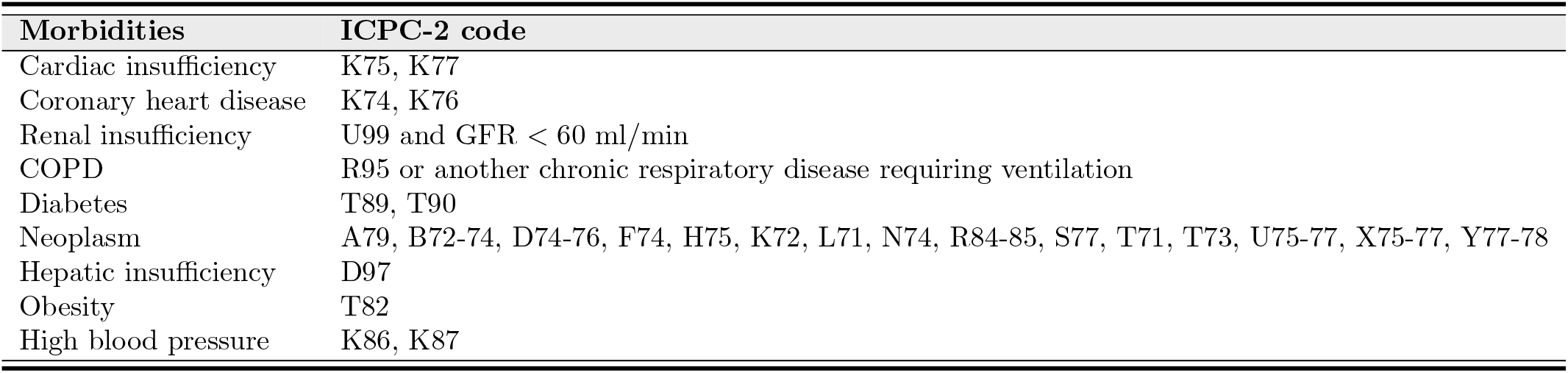
ICPC-2 codes for morbidities specified in the Portuguese vaccination plan.

| Morbidities | ICPC-2 code |
| --- | --- |
| Cardiac insufficiency | K75, K77 |
| Coronary heart disease | K74, K76 |
| Renal insufficiency | U99 and GFR < 60 ml/min |
| COPD | R95 or another chronic respiratory disease requiring ventilation |
| Diabetes | T89, T90 |
| Neoplasm | A79, B72-74, D74-76, F74, H75, K72, L71, N74, R84-85, S77, T71, T73, U75-77, X75-77, Y77-78 |
| Hepatic insufficiency | D97 |
| Obesity | T82 |
| High blood pressure | K86, K87 |

**Table S2.** Summary of the model parameters.

| Description (unit) | Notation | Reference |
| --- | --- | --- |
| **Constant parameters** |  |  |
| Latent period (days) | $1/\alpha$ | Estimated |
| Infectious period (days) | $1/\gamma$ | Estimated |
| Over-dispersion parameter for the NegBinom distribution for hospitalizations | $\phi$ | Estimated |
| Initial fraction of infected persons | $\theta$ | Estimated |
| Probability of transmission per contact | $\epsilon$ | Estimated |
| **Age-specific parameters\*** |  |  |
| Force of infection for unvaccinated and vaccinated persons (1/day) | $\lambda_k(t), \lambda_k^V(t)$ | Eqs. (3) and (4) |
| Contact rate for unvaccinated persons (1/day) | $c_{kl}(t)$ | Estimated, see Table S4 |
| Contact rate for vaccinated persons (1/day) | $c_{kl}^V(t)$ | Assumed |
| Hospitalization rate (1/day) | $\nu_k$ | Estimated |
| Susceptibility of age group $k$ relative to age group $n = 10$ | $\beta_k$ | Estimated |
| Population size of age group $k$ | $N_k$ | [66] |
| **Vaccination parameters\*** |  |  |
| Vaccination rate (1/day) | $r_k$ | Calculated from Table 1 and Figure 4 a |
| Vaccine efficacy in reducing susceptibility | $VE_S$ | 94% (optimistic), 55% (pessimistic) [15, 18–23, 26] |
| Vaccine efficacy in reducing infectivity | $VE_I$ | 0% (optimistic), 0% (pessimistic) [15, 18–23, 26] |
| Vaccine efficacy in reducing hospitalization rate | $VE_H$ | 67% (optimistic), 0% (pessimistic) [15, 18–23, 26] |

\*Indices $k$ and $l$ denote the age groups $k, l = 1, \dots, n$ , where $n = 10$ is the number of age groups.

**Table S3.**
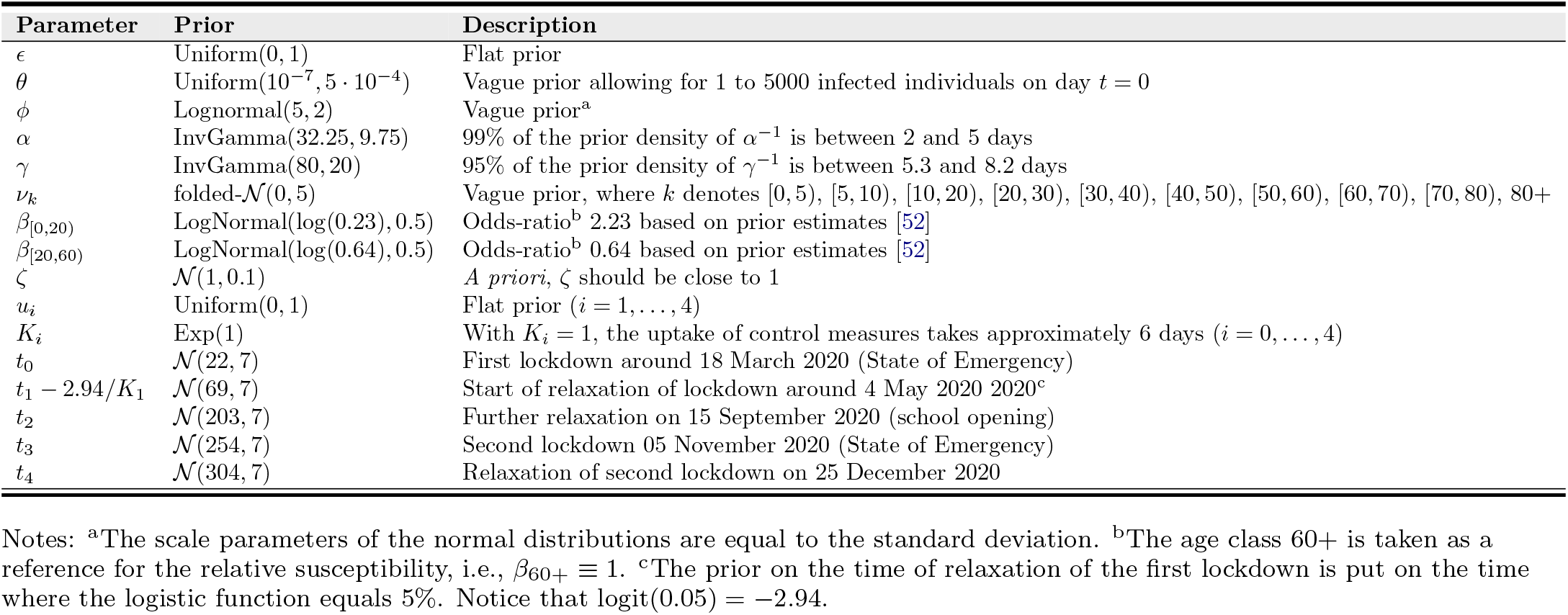
Prior distribution of the statistical model.

**Table S4.** Parameters describing contact structure.

| Description (unit) | Notation* | Reference |
| --- | --- | --- |
| **Contact rates (1/day)** |  |  |
| Baseline (pre-pandemic) | $b_{kl}$ | [67] |
| After the first lockdown | $\zeta a_{kl}$ | $\zeta$ estimated, $a_{kl}$ inferred using [68] |
| After the first relaxation | $u_1 b_{kl} + (1 - u_1) \zeta a_{kl}$ | Estimated |
| After the second relaxation due to school opening | $u_2 b_{kl} + (1 - u_2) \zeta a_{kl}$ | Estimated |
| After the second lockdown | $u_3 b_{kl} + (1 - u_3) \zeta a_{kl}$ | Estimated |
| After the relaxation due to winter holidays | $u_4 b_{kl} + (1 - u_4) \zeta a_{kl}$ | Estimated |
| After the third lockdown | $\zeta a_{kl}$ | Assumed |
| After first relaxation during the vaccination rollout (Scenario 1) | $b_{kl}$ | Assumed |
| After first relaxation during the vaccination rollout (Scenario 2) | $u_2 b_{kl} + (1 - u_2) \zeta a_{kl}$ | Assumed |
| After first relaxation during the vaccination rollout (Scenario 3) | $u_1 b_{kl} + (1 - u_1) \zeta a_{kl}$ | Assumed |
| After first, second, third relaxation during the vaccination rollout (Scenario 4) | Matrices for Scenario 3, 2, 1 | Assumed |
| **Mid-point time of the logistic function (days)** |  |  |
| Introduction of the first lockdown | $t_0$ | Estimated |
| Relaxation after the first lockdown | $t_1$ | Estimated |
| Second relaxation due to school opening | $t_2$ | Estimated |
| Introduction of the second lockdown | $t_3$ | Estimated |
| Relaxation due to winter holidays | $t_4$ | Estimated |
| Introduction of the third lockdown | $t_5$ | 28 January 2021, Assumed |
| First relaxation during the vaccination rollout | $t_6$ | 1 April 2021, Assumed |
| Second relaxation during the vaccination rollout | $t_7$ | 1 June 2021, Assumed |
| Third relaxation during the vaccination rollout | $t_8$ | 1 October 2021 (main analyses), 1 August (sensitivity analyses), Assumed |
| **Slope of the logistic function (1/day)** |  |  |
| Introduction of the first lockdown | $K_0$ | Estimated |
| Relaxation after the first lockdown | $K_1$ | Estimated |
| Second relaxation due to school opening | $K_2$ | Estimated |
| Introduction of the second lockdown | $K_3$ | Estimated |
| Relaxation due to winter holidays | $K_4$ | Estimated |
| Introduction of the third lockdown | $K_0$ | Assumed |
| First relaxation during the vaccination rollout | $K_1$ | Assumed |
| Second relaxation during the vaccination rollout | $K_1$ | Assumed |
| Third relaxation during the vaccination rollout | $K_1$ | Assumed |
| **Proportion of time a person behaves as before the pandemic** |  |  |
| Relaxation after the first lockdown | $u_1$ | Estimated |
| Second relaxation due to school opening | $u_2$ | Estimated |
| Introduction of the second lockdown | $u_3$ | Estimated |
| Relaxation due to winter holidays | $u_4$ | Estimated |

\*Indices $k$ and $l$ denote the age groups $k, l = 1, \dots, n$ , where $n = 10$ is the number of age groups.

